# Association of Polygenic Risk Scores for Neurodevelopmental Traits and Psychiatric Conditions with Incontinence and Constipation in Children and Young People

**DOI:** 10.1101/2024.11.29.24318188

**Authors:** Oliver Bastiani, Christina Dardani, Kimberley Burrows, Jane Hvarregaard Christensen, Carol Joinson

## Abstract

Observational studies report prospective associations of neurodevelopmental and psychiatric traits with paediatric incontinence and constipation, but unmeasured and residual confounding may limit observational measures. Here, we use a prospective birth cohort study to investigate whether common variant genetic liability for a range of neurodevelopmental and psychiatric conditions are associated with paediatric incontinence and constipation. We used data from 7,857 participants from the Avon Longitudinal Study of Parents and Children (ALSPAC) with data on genotype, incontinence, and constipation, and calculated Polygenic risk scores (PRS) for neurodevelopmental traits (ADHD, autism, intelligence) and psychiatric conditions (anxiety, depression, and OCD). Incontinence subtypes (daytime urinary incontinence [DUI], enuresis [any bedwetting and enuresis subtypes: monosymptomatic, non-monosymptomatic], faecal incontinence), and constipation, were assessed by parental reports at age 9 years and self-reports at age 14. PRS for ADHD (OR=1.14, 95% CI, 1.01-1.29, unadjusted *p*=0.040) and depression (OR=1.09, 95% CI, 1.00–1.20, unadjusted *p*=0.063) were associated with DUI at age 9. PRS for autism (OR=1.19, 95% CI, 1.02-1.41, unadjusted *p*=0.032) and intelligence (OR=1.17, 95%, 0.99-1.38, unadjusted *p*=.06l) were associated with DUI at age 14. PRS for ADHD (OR=1.13, 95% CI, 1.03-1.24, unadjusted *p*=0.008) were associated with constipation at age 9. Within enuresis subtypes, PRS for autism were associated with MNE at age 9 (OR=1.15, 95% CI, 1.03–1.28, unadjusted *p*=0.012), but not NMNE (OR=0.93, 95% CI, 0.79–1.18, unadjusted *p*=0.335). No associations survived false discovery rate adjustment. The findings add to existing evidence that common variant genetic liability for neurodevelopmental traits and psychiatric conditions could be associated with paediatric incontinence and constipation.

**Key points:** *Question:* Are common variant genetic liabilities for neurodevelopmental and psychiatric conditions associated with paediatric incontinence and constipation in a population-based cohort?

*Findings:* We found some evidence that polygenic risk scores (PRS) for ADHD, autism, intelligence, and depression may be associated with daytime urinary incontinence. PRS for ADHD were also associated with constipation and enuresis and PRS for autism and depression were weakly associated with constipation. None of the associations survived adjustment for false discovery rate.

*Meaning:* Common variant genetic liabilities for ADHD, autism, intelligence, and depression could be risk factors for developing paediatric incontinence and constipation.

## Introduction

Bedwetting (enuresis), daytime urinary incontinence (DUI), soiling (fecal incontinence), and constipation are common problems in childhood and they negatively impact children and their families(1, 2). Children and young people who experience incontinence are at increased risk of peer victimisation, depressive symptoms, poor self-image, and adverse mental health outcomes in later adolescence(3, 4). Amongst 7-year-olds, the prevalence of bedwetting, DUI, and soiling are estimated at 15%, 7.8% and 6.8% respectively(5–7). The prevalence of incontinence decreases with age (e.g., DUI prevalence decreases to around 4.9% at age 9 and 2.9% at age 14(8, 9)) whilst the prevalence of constipation is more stable across childhood (estimated median prevalence of 8.9%(10)). DUI and soiling are common consequences of chronic constipation(11).

Risk factors for incontinence and constipation are multifactorial and involve a complex interaction of genetic, biological, neurological, psychosocial, and environmental factors. Cross-sectional studies suggest that incontinence and constipation are more common in children with neurodevelopmental traits (e.g. attention deficit hyperactivity disorder [ADHD] symptoms, autistic traits, lower IQ) and psychiatric conditions (e.g. depression, anxiety, and obsessive compulsive disorder [OCD])(12, 13). There is also evidence that emotional and behavioural problems (e.g. ADHD symptoms, anxiety, low mood) are prospectively associated with incontinence and constipation(14).

Prospective studies provide evidence of the direction of associations but are limited by unmeasured and residual confounding(15). Observational studies that use parental reports of their child’s neurodevelopmental/psychiatric symptoms are also limited by measurement error because questionnaire assessments could misclassify cases and non-cases(16), and could be biased by maternal reports(17). The increasing availability of genome-wide association studies (GWASs) of neurodevelopmental traits and psychiatric conditions enables the derivation of polygenic risk scores (PRS)(18), which estimate an individual’s underlying common variant genetic liability to a complex trait(18). PRSs are typically not associated with environmental confounders at a population level, which can bias observational studies, and they can potentially minimize measurement error that also affects observational studies.

The current study, based on data from a large UK birth cohort, investigates if common variant genetic liability for neurodevelopmental traits and psychiatric conditions is associated with different subtypes of incontinence (bedwetting, DUI, and soiling) and constipation in childhood (parent-reported at 9 years) and adolescence (self-reported at 14 years). We hypothesized that common variant genetic liability for neurodevelopmental traits and psychiatric conditions would be associated with an increase in the odds of incontinence and constipation. We also examined associations between the PRS and different subtypes of enuresis because there is evidence that psychiatric conditions are more strongly associated with non-monosymptomatic enuresis (bedwetting with daytime lower urinary tract symptoms [LUTS]) than the monosymptomatic subtype (bedwetting without daytime LUTS)(19).

## Methods

### Study Population

Data were obtained from the Avon Longitudinal Study of Parents and Children (ALSPAC)(20–22). ALSPAC is a prospective, population-based birth cohort study that recruited pregnant women resident in Avon, UK with expected dates of delivery between 1st April 1991 and 31st December 1992. The initial number of pregnancies enrolled was 14,541 resulting in 14,062 live births and 13,988 children who were alive at 1 year of age. When the oldest children were approximately 7 years of age, an attempt was made to bolster the initial sample with eligible cases who had failed to join the study originally resulting in an additional 913 children being enrolled. The total sample size for analyses using any data collected after the age of seven is therefore 15,447 pregnancies, resulting in 15,658 foetuses. Of these 14,901 children were alive at 1 year of age. The phases of enrolment are described in more detail in the cohort profile paper and its update. The study website (www.bristol.ac.uk/alspac) contains details of all the data that is available through a fully searchable data dictionary and variable search tool: http://www.bristol.ac.uk/alspac/researchers/our-data/. Ethical approval for the study was obtained from the ALSPAC Ethics and Law Committee and the Local Research Ethics Committees. Informed consent for the use of data collected via questionnaires and clinics was obtained from participants following the recommendations of the ALSPAC Ethics and Law Committee at the time. Consent for biological samples has been collected in accordance with the Human Tissue Act (2004). The current study included all children with genotype data and data for incontinence and/or constipation, at 9 and 14 years. Sample derivation is shown in Figure 1 and sample characteristics are shown in table 1.

**Figure 1.**
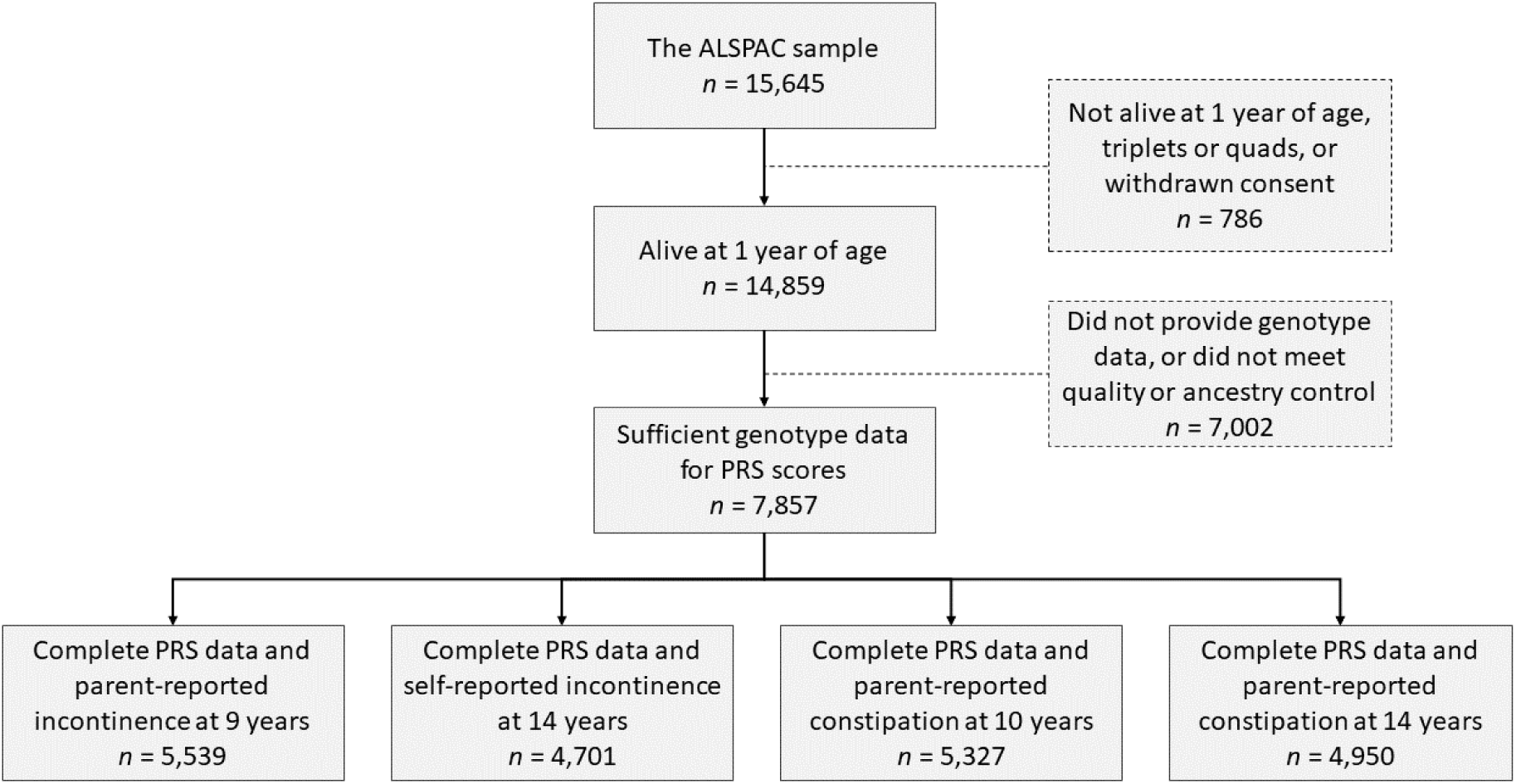
Flow diagram depicting eligible samples in the present study

**Table 1.** Distribution of characteristics across the samples. Due to questionnaire completion rates, and attrition with age, sample sizes differed between analyses (Figure 1), the largest samples for each analysis are presented here. Socioeconomic factors were derived from maternal questionnaire responses during pregnancy (between 8 and 32 weeks gestation) including: occupational social class (using the 1991 British Office of Population and Census Statistics classification and dichotomised into ‘non- manual’ and ‘manual’); maternal education (‘A-level or greater’, ‘O level’, and ‘Certificate of Secondary (CSE) Education/vocational qualification/none’); home ownership (‘mortgaged/owned/private renter’ and ‘rented from council or housing association/non- homeowner’). Parity was coded as whether study child is 1st/2nd/3rd child or greater)

|  | Starting Sample<br>(N ≤ 15,645) | Sample with PRS<br>Data (N ≤ 7,857) | PRS data and<br>Incontinence at<br>age 9 (N ≤ 5,539) | PRS data and<br>Incontinence at age<br>14 (N ≤ 4,139) |
| --- | --- | --- | --- | --- |
| Child's Sex (ref: Female) | 7,683 (51.1%) | 4,014 (51.2%) | 2,804 (50.6%) | 2,149 (45.8%) |
| Parental Social Class (ref: non-manual) | 2,245 (19.4%) | 1,066 (15.9%) | 647 (13.0%) | 522 (12.3%) |
| Maternal Education (ref: A-level or greater) |  |  |  |  |
| CSE/vocational/none | 3,753 (30.1%) | 1,730 (24.5%) | 1,017 (19.6%) | 819 (18.6%) |
| O-Level | 4,318 (34.6%) | 2,455 (34.8%) | 1,822 (35.1%) | 1,496 (33.4%) |
| Home Ownership Status |  |  |  |  |
| (ref: Mortgaged/ owned/private renter) |  |  |  |  |
| Rented from council or housing<br>association/non-homeowner | 3,615 (26.8%) | 1,480 (20.5%) | 817 (15.7%) | 643 (14.6%) |
| Parity (ref: 0 pregnancies) |  |  |  |  |
| 1 pregnancy | 4,575 (29.7%) | 2,609 (33.6%) | 1,889 (34.4%) | 1,593 (34.2%) |
| 2+ pregnancies | 4,971 (32.3%) | 1,951 (25.1%) | 1,208 (22.0%) | 1,007 (21.6%) |

## Measures

### Incontinence and constipation

Mothers completed a questionnaire when their child was age 9 including the following questions on incontinence: “How often does your child wet the bed at night; wet his/herself during the day; dirty his/her pants during the day?”. Response options included “Never”, “Occasional accidents less than once a week”, “About once a week”, “Two to five times a week”, “Nearly every day” and “More than once a day”. We derived binary variables separately for bedwetting, DUI and soiling to indicate the presence of any incontinence (≥occasional) versus none.

Constipation was assessed by parental questionnaires at 10 and 14 years. The questionnaires asked, “Has your child had constipation in the past 12 months?”. Options included “Yes, saw a doctor”, “Yes, did not see a doctor”, and “No, did not have”. We derived binary variables for each age by categorising the first two options as cases and the third option as non-cases.

### Enuresis subtypes

We categorized children into probable cases of non-monosymptomatic enuresis (NMNE: bedwetting in the presence of DUI and/or urgency) and monosymptomatic enuresis (MNE: bedwetting in the absence of DUI and urgency). We examined two binary variables indicating (i) the presence versus absence of NMNE and (ii) the presence versus absence of MNE. Urgency was assessed via a parent-completed questionnaire at age 9 and a child-completed questionnaire at age 14. Parents were asked “Does he/she have to dash to the toilet quickly when he/she realises he/she needs to go?”. Options included “Yes, had to go straight away”, “Can hold for a short time (less than 5 minutes)”, and “Can hold longer than 5 minutes”. We categorized the first option as urinary urgency cases, and the remaining options as non-cases. At age 14, study children were asked “Over the last 2 weeks, how often have you had a sudden feeling you needed a wee and had to dash to the toilet?”. Response options included ‘never’, ‘a few times’, ‘quite often’, ‘a lot’. We categorized the second two options as urinary urgency cases and the first two options as non-cases.

### PRS generation for neurodevelopmental and psychiatric conditions

#### Discovery Sample

PRSs were calculated using the latest publicly available GWAS data on autism (Ncases = 18,381; Ncontrols = 27,969)(23), ADHD (Ncases = 38,691; Ncontrols = 186,843)(24), anxiety (Ncases = 18,186; Ncontrols = 17,310)(25), depression (Ncases = 371,184; Ncontrols = 978,703)(26), OCD (Ncases = 2,688; Ncontrols = 7,037)(27), and intelligence (N = 269,867)(28).

#### Target Sample

After quality control and excluding participants who had withdrawn consent, genetic data were available for 7,857 children of European ancestry. For the full description of how the genotype data was obtained, please see Supplementary Methods 1.

#### PRS Estimation in the Target Sample

PRS were calculated using PLINK v.1.9, applying the method described by the Psychiatric Genomics Consortium (PGC). This approach is consistent with current guidelines for the use of PRSs in the context of psychiatric conditions(29). We removed single nucleotide polymorphisms (SNPs) with mismatching alleles between the discovery and target dataset.

The Major Histocompatibility Complex (MHC) region was removed (25–34 Mb), except for one SNP representing the strongest signal within the region. Using ALSPAC data as the reference panel, SNPs were LD clumped with an r^2^ of 0.25 and a physical distance threshold of 500 kB. The optimal *p*-value threshold for PRS is dependent on discovery and target sample sizes, as well as SNP inclusion parameters (for example, r^2^). Thus, PRS for each participant were calculated across 13 *p*-value thresholds (P < 5.0 × 10^−8^ to P < 0.5), standardized by subtracting the mean and dividing by the standard deviation. We defined PRS corresponding to *p*-value threshold 0.05 as our primary exposure, based on previous ALSPAC studies(30).

## Data analysis

### Primary Analyses

We investigated the association between the PRS for six neurodevelopmental traits and psychiatric conditions and incontinence (DUI, bedwetting, and soiling) and constipation occurring at ages 9 and 14. We used logistic regression analyses to test the associations separately for each PRS with incontinence and constipation in each age group with adjustment for sex and the first 10 principal components in ALSPAC. We calculated false discovery rate (FDR) adjusted *p*-value thresholds to control for Type-1 error across 48 tests in total(31).

### Analysis of enuresis subtypes

We examined the associations between each PRS exposure and the enuresis subtypes (probable NMNE and MNE), at ages 9 and 14 years. We used logistic regression analysis and adjusted for sex and the first 10 principal components in ALSPAC.

## Results

The sample sizes for the analyses varied between incontinence subtypes due to data availability. Sample sizes were smaller at age 14 (Figure 1). Compared with the original ALSPAC sample, the participants in the analyses at ages 9 and 14 had a higher proportion of indicators of higher socioeconomic status and a lower proportion of mothers with two or more pregnancies in addition to the study child (Table 1). Figure 2 shows the prevalence of each incontinence/constipation outcome at each age.

**Figure 2.**
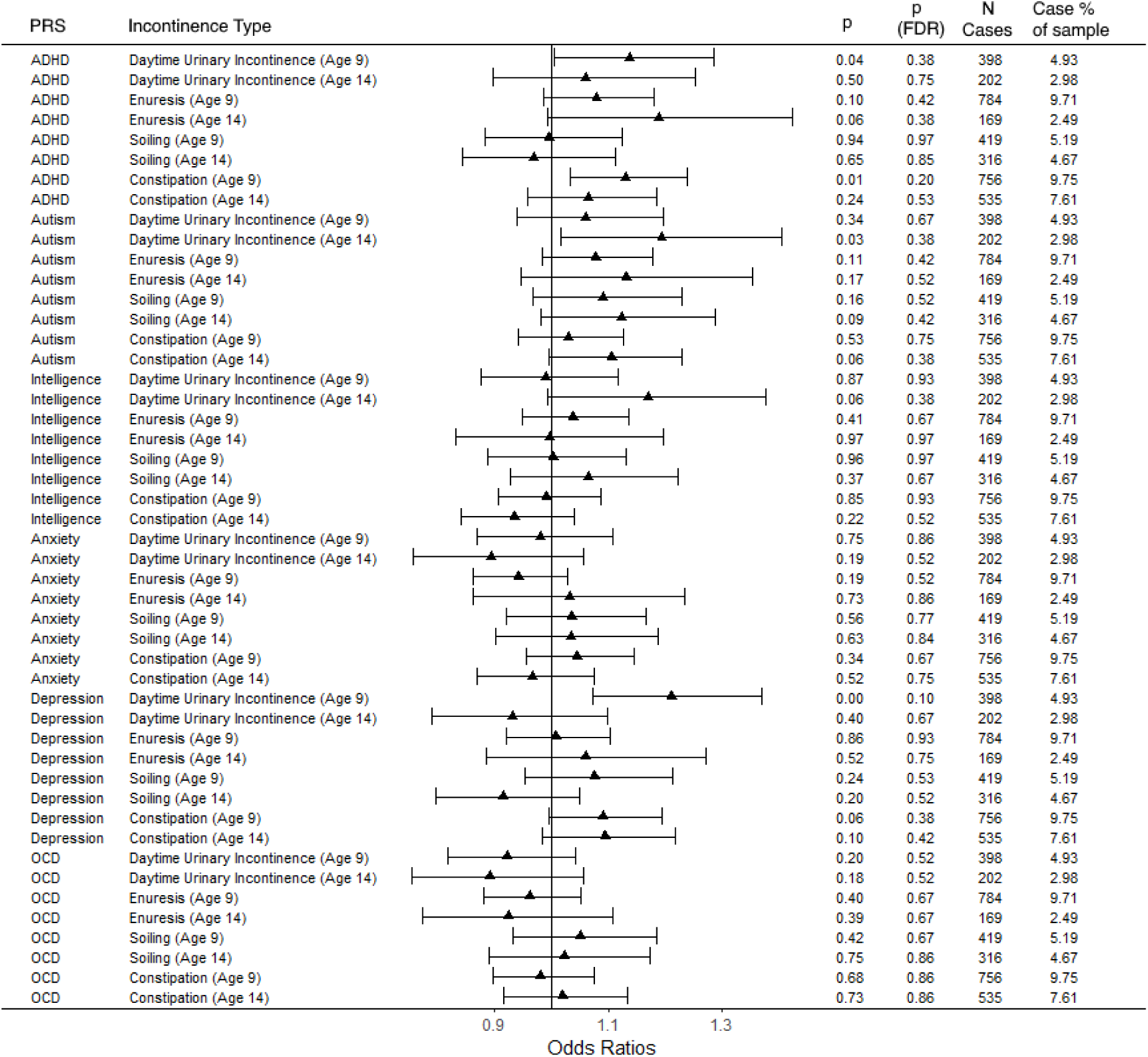
Associations between polygenic risk scores at the .05 *p*-value threshold and incontinence types in the ALSPAC cohort adjusted for sex and the first 10 principal components in ALSPAC. Results present the odds ratios and 95% CIs, unadjusted *p*-values, FDR-corrected *p*-values, and the frequency of incontinence subtype cases in each analysis. Due to space constraints and clarity of our primary findings, Supplementary Figure 6 presents the effect estimates (ORs) and 95% confidence intervals of these associations. The supplement also includes analyses between incontinence and constipation and all PRSs for SNP-inclusion significance levels ranging from p<.05 to < 5*10^-8^ (S1-S13).

### PRS for neurodevelopmental traits

Figure 2 shows odds ratios (OR), *p*-values, and sample sizes for the analysis of PRS scores at the p<0.05 SNP-inclusion significance level. There was evidence of associations between the ADHD PRS and DUI at age 9 (OR, 1.14; 95% CI, 1.01-1.29, unadjusted *p* =.040), and constipation at age 9 (OR, 1.13; 95% CI, 1.03-1.24, unadjusted *p* =.008). There was weaker evidence of associations with enuresis at ages 9 (OR, 1.08; 95% CI, 0.99–1.18, unadjusted *p* =.095), and 14 (OR, 1.19; 95% CI, 0.99–1.43, unadjusted *p* =.059).

The autism PRS was associated with DUI at age 14 (OR, 1.19; 95% CI, 1.02-1.41, unadjusted *p* =.032), but not at age 9 (OR, 1.06; 95% CI, 0.94–1.20, unadjusted *p* =.340) and weakly associated with constipation at age 14 (OR, 1.11; 95% CI, 0.99–1.23, unadjusted *p* =.062). There was little evidence of associations between the PRS for intelligence and incontinence/constipation, except for weak evidence of an association with DUI at age 14 (OR, 1.17; 95%, 0.99-1.38, unadjusted *p* =.06l).

None of the associations between the PRS for neurodevelopmental traits and incontinence/constipation survived FDR adjustments.

### PRS for psychiatric conditions

There was evidence of an association between the depression PRS and DUI at age 9 (OR, 1.21; 95% CI, 1.07–1.37, unadjusted *p* <.001), and weak evidence of an association with constipation at age 9 (OR, 1.09; 95% CI, 1.00–1.20, unadjusted *p* =.063). There was no evidence that the PRS for anxiety or OCD were associated with incontinence or constipation.

None of the associations between the PRS for psychiatric conditions and incontinence/constipation survived FDR adjustments.

Associations of the PRS for neurodevelopmental traits and psychiatric conditions with probable monosymptomatic and non-monosymptomatic enuresis

There was little evidence of differential associations between the PRS for neurodevelopmental traits or psychiatric conditions and the enuresis subtypes, and no associations survived FDR adjustments. The exception was the autism PRS, which was associated with probable monosymptomatic enuresis at age 9 (OR, 1.15; 95% CI, 1.03–1.28, unadjusted *p* =.012), but not with non-monosymptomatic enuresis (OR, 0.93; 95% CI, 0.79– 1.18, unadjusted *p* =.335). However, the 95% confidence intervals overlapped.

Contrary to our hypothesis, increased common variant genetic liability for OCD was associated with reduced odds of non-monosymptomatic enuresis at age 9 (OR, 0.80; 95% CI, 0.68–0.93, unadjusted *p* =.005). Figure 3 shows the odds ratios, *p*-values, and sample sizes for the analysis.

**Figure 3.**
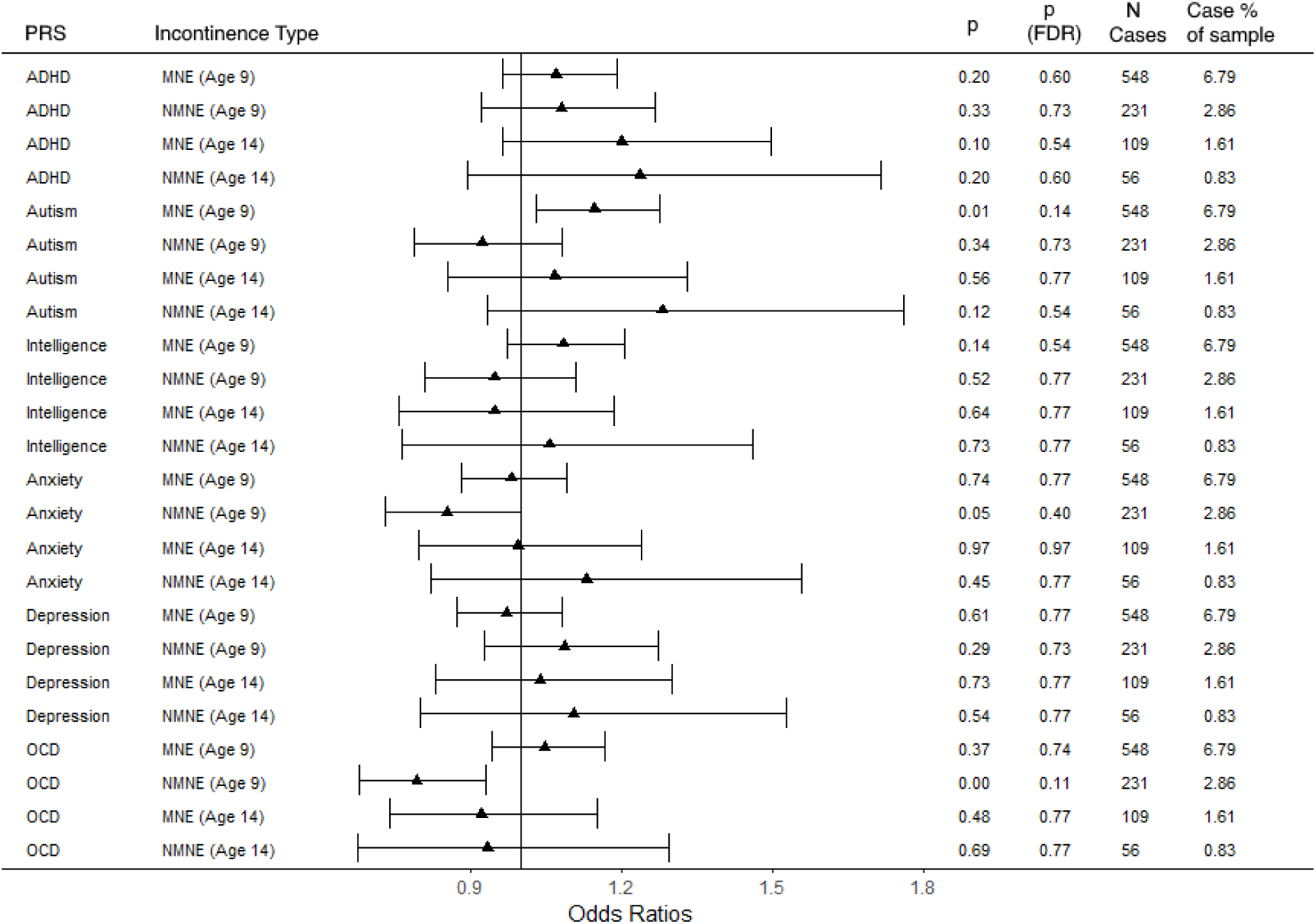
Associations between polygenic risk scores at the .05 significance level and monosymptomatic (MNE) and non-monosymptomatic (NMNE) enuresis in the ALSPAC cohort adjusted for sex and the first 10 principal components in ALSPAC. Results present the odds ratios and 95% CIs, unadjusted *p*-values, FDR-corrected *p*-value, and the frequency of monosymptomatic or non-monosymptomatic enuresis cases in each analysis.

## Discussion

In this study we examined associations between common variant genetic liabilities for neurodevelopmental traits and psychiatric conditions, and paediatric incontinence and constipation. To our knowledge, this is the first birth cohort study to investigate these associations. We found evidence of associations between the ADHD PRS and incontinence (daytime wetting at age 9, enuresis at ages 9 and 14) and constipation (age 9) and evidence of an association between the PRS for autism and incontinence/constipation. A previous GWAS of enuresis conducted in the Danish iPSYCH2012 case-cohort sample explored the associations between PRS for ADHD/autism and enuresis in a subset of participants with no reported ADHD/autism(32) and found an association between the ADHD PRS and enuresis, but did not find an association between the autism PRS and enuresis. Furthermore, a GWAS of DUI, conducted in the Danish iPSYCH2015 cohort sample(33), found associations between PRS for ADHD and DUI. Enuresis/DUI cases in these previous studies were identified based on diagnostic codes and redeemed prescriptions for enuresis/DUI medications whilst our study is not restricted to children who met clinical diagnostic criteria. Our findings therefore apply to children in the wider community with a range of severities of incontinence/constipation, rather than only children with clinical diagnoses.

Additionally, the enuresis GWAS did not distinguish between enuresis subtypes. When we examined evidence for differential associations between the PRS and the different enuresis subtypes, we found weak evidence that the autism PRS was more strongly associated with monosymptomatic enuresis, than non-monosymptomatic enuresis, at age 9. More research must investigate the association between ASD and enuresis subtypes because no previous studies have examined this(13).

Compared with existing observational studies, our study found weaker (or no) contribution of genetic liability to neurodevelopmental traits/psychiatric conditions on incontinence and constipation(12, 34–37). Observational studies have found prospective associations between anxiety and emotional/behavioural problems and paediatric urinary incontinence(14, 38, 39). These different findings may be due to confounding in observational studies and their reliance on questionnaire-based measures. An alternative explanation is that the PRS explain only a small proportion of the genetic variance in psychiatric conditions, and rely on common genetic variation, being unable to capture fully the genetic component of these conditions (i.e., common and rare variation)(18).

There was weak evidence of associations between the depression PRS and DUI (age 9 only) and constipation. The PRS for intelligence was also weakly associated with DUI at age 14. We found no evidence of associations between the PRS for anxiety and OCD and incontinence/constipation. Contrary to our hypothesis, the PRS for OCD was associated with reduced odds of non-monosymptomatic enuresis (at age 9 only). Observational studies have reported conflicting results regarding the association between obsessive-compulsive traits and enuresis(40), but did not distinguish between enuresis subtypes. A birth cohort study found evidence of stronger associations between non-mono-, compared with monosymptomatic, enuresis and psychiatric conditions(19). Further research with larger samples is needed to examine if neurodevelopmental/psychiatric traits are differentially associated with the enuresis subtypes.

A strength of our study is the use of PRS for neurodevelopmental traits/psychiatric conditions. PRS are typically not associated with factors which can confound observational studies because genetic variants are randomly assigned at conception. Previous studies that examined associations between neurodevelopmental/psychiatric traits and incontinence/constipation relied on maternal reports of neurodevelopmental/psychiatric traits which could differ from the child’s self-reports(17), and may have been biased by the mothers’ own psychiatric states or mood(41). We used data from both parent and child assessments of incontinence in childhood and adolescence (9 and 14 years respectively). PRS may also capture traits that are not phenotypically expressed until later stages of the condition, thus, would be missed on typical questionnaire assessments(42). The GWAS data used to generate our PRS were the latest and most highly powered. Our analysis also included a range of common variant genetic liability for traits, rather than being restricted to only those who met clinical diagnostic criteria for conditions. We also examined different subtypes of enuresis (probable mono-and non-monosymptomatic), whilst other paediatric incontinence studies rarely discriminate between these subtypes.

The study has several limitations which should be considered when interpreting the findings. PRS only measure common variant genetic liability, so will miss instances where genetic and environmental risk factors are distinct, which can occur in psychiatric conditions, such as depression(43, 44). However, PRSs exclude other, potentially confounding, factors related to the phenotypical expression of traits that may influence their relationship with incontinence/constipation. Furthermore, no associations survived FDR adjustment. However, our conclusions are not purely based on *p*-value thresholds (e.g. p<0.05) to determine statistical significance, instead, we consider the effect estimates alongside the strength of evidence indicated by the *p*-values and confidence intervals in line with current recommendations(45, 46).

The GWAS samples used to inform our PRS consisted mainly of adult European ancestry samples, so our findings may not generalize beyond these populations. Additionally, some conditions, such as depression, are phenotypically different for adults and children(16), therefore, PRS derived from adult samples may not be reliable measures of paediatric psychiatric conditions.

Attrition bias due to selective dropout is a potential limitation because the participants that comprised the sample that provided genetic data in addition to incontinence data were more socioeconomically advantaged compared with the original ALSPAC cohort. However, studies have reported that incontinence and constipation are only weakly socially patterned in the ALSPAC cohort(39, 47).

## Conclusion

We have found some evidence suggesting that common variant genetic liabilities for neurodevelopmental and psychiatric conditions may be associated with paediatric incontinence and constipation. Early assessment and treatment of bladder and bowel issues should be considered for children with neurodevelopmental/psychiatric traits to reduce the risk of chronic incontinence and constipation.

Supplementary information is available at Molecular Psychiatry’s website

## Data Availability

Due to ALSPAC regulations, we cannot provide access to this data but if others wish to access this data they may contact the involved cohort study (ALSPAC) at https://www.bristol.ac.uk/alspac/researchers/access/, where they may apply to use this data also.

## Acknowledgements

We are extremely grateful to all the families who took part in this study, the midwives for their help in recruiting them, and the whole ALSPAC team, which includes interviewers, computer and laboratory technicians, clerical workers, research scientists, volunteers, managers, receptionists and nurses.

