## Supplementary Methods and Tables for "Association of Polygenic Risk Scores for Neurodevelopmental Traits and Psychiatric Conditions with Incontinence and Constipation in Children and Young People"

Online Resource

Supplementary Information Contents:

Supplementary Methods 1. ALSPAC Genotyping

Supplementary Figure 1. Coefficients and 95% confidence intervals for the analysis of PRS Scores at the $p < 0.5$ SNP-inclusion significance level (S1)

Supplementary Figure 2. Coefficients and 95% confidence intervals for the analysis of PRS Scores at the $p < 0.4$ SNP-inclusion significance level (S2)

Supplementary Figure 3. Coefficients and 95% confidence intervals for the analysis of PRS Scores at the $p < 0.3$ SNP-inclusion significance level (S3)

Supplementary Figure 4. Coefficients and 95% confidence intervals for the analysis of PRS Scores at the $p < 0.2$ SNP-inclusion significance level (S4)

Supplementary Figure 5. Coefficients and 95% confidence intervals for the analysis of PRS Scores at the $p < 0.1$ SNP-inclusion significance level (S5)

Supplementary Figure 6. Coefficients and 95% confidence intervals for the analysis of PRS Scores at the $p < 0.05$ SNP-inclusion significance level (S6)

Supplementary Figure 7. Coefficients and 95% confidence intervals for the analysis of PRS Scores at the $p < 0.01$ SNP-inclusion significance level (S7)

Supplementary Figure 8. Coefficients and 95% confidence intervals for the analysis of PRS Scores at the $p < 0.001$ SNP-inclusion significance level (S8)

Supplementary Figure 9. Coefficients and 95% confidence intervals for the analysis of PRS Scores at the $p < 0.0001$ SNP-inclusion significance level (S9)

Supplementary Figure 10. Coefficients and 95% confidence intervals for the analysis of PRS Scores at the $p < 0.00001$ SNP-inclusion significance level (S10)

Supplementary Figure 11. Coefficients and 95% confidence intervals for the analysis of PRS Scores at the $p < 0.000001$ SNP-inclusion significance level (S11)

Supplementary Figure 12. Coefficients and 95% confidence intervals for the analysis of PRS Scores at the $p < 0.0000001$ SNP-inclusion significance level (S12)

Supplementary Figure 13. Coefficients and 95% confidence intervals for the analysis of PRS Scores at the $p < 0.00000005$ SNP-inclusion significance level (S13)

Supplementary Methods 1. ALSPAC Genotyping

A total of 9,912 ALSPAC children were genotyped on the Illumina HumanHap550 quad chip genotyping platforms by 23andme subcontracting the Wellcome Trust Sanger Institute, Cambridge, UK, and the Laboratory Corporation of America, Burlington, NC, United States.

PLINK v1.07 was used for quality control filtering. Specifically, individuals were excluded on the basis of the following filters: (1) gender mismatches; (2) undetermined X chromosome heterozygosity; (3) over 3% missingness (children); over 5% missingness (mothers); (4) evidence of cryptic relatedness (>10% of shared alleles identical by descent in children and >12.5% of shared alleles identical by descent in mothers); (5) non-European ancestry, assessed by multidimensional scaling analysis compared with HapMap 2 individuals. SNPs were excluded on the basis of the following filters: (1) minor allele frequency < 1%; (2) call rate < 95%, (3) Hardy–Weinberg equilibrium (HWE) $P < 5.0 \times 10{-}7$. Maternal and offspring genotype data were combined and imputed using Impute v.2.2.2 against 1000 Genomes reference panel (v.1, phase 3, December 2013 release).

Supplementary Figure 1. Coefficients and 95% confidence intervals for the analysis of PRS Scores at the p < 0.5 SNP-inclusion significance level (S1)

| PRS | Incontinence Type | Odds Ratios (95% CI) | *p* |
|---|---|---|---|
| ADHD | Daytime Urinary Incontinence (Age 9) | 1.08 (0.95-1.22) | 0.25 |
| ADHD | Daytime Urinary Incontinence (Age 14) | 1.04 (0.88-1.23) | 0.61 |
| ADHD | Enuresis (Age 9) | 1.09 (0.99-1.19) | 0.07 |
| ADHD | Enuresis (Age 14) | 1.24 (1.04-1.48) | 0.02 |
| ADHD | Soiling (Age 9) | 1.01 (0.89-1.14) | 0.87 |
| ADHD | Soiling (Age 14) | 0.98 (0.85-1.12) | 0.75 |
| ADHD | Constipation (Age 9) | 1.15 (1.05-1.26) | 0.00 |
| ADHD | Constipation (Age 14) | 1.10 (0.98-1.22) | 0.09 |
| Autism | Daytime Urinary Incontinence (Age 9) | 1.14 (1.01-1.28) | 0.04 |
| Autism | Daytime Urinary Incontinence (Age 14) | 1.17 (0.99-1.39) | 0.06 |
| Autism | Enuresis (Age 9) | 1.06 (0.96-1.16) | 0.24 |
| Autism | Enuresis (Age 14) | 1.15 (0.96-1.38) | 0.13 |
| Autism | Soiling (Age 9) | 1.08 (0.96-1.22) | 0.22 |
| Autism | Soiling (Age 14) | 1.15 (1.00-1.32) | 0.05 |
| Autism | Constipation (Age 9) | 1.05 (0.96-1.14) | 0.33 |
| Autism | Constipation (Age 14) | 1.08 (0.98-1.21) | 0.13 |
| Intelligence | Daytime Urinary Incontinence (Age 9) | 0.98 (0.87-1.11) | 0.80 |
| Intelligence | Daytime Urinary Incontinence (Age 14) | 1.22 (1.04-1.44) | 0.02 |
| Intelligence | Enuresis (Age 9) | 1.03 (0.94-1.13) | 0.51 |
| Intelligence | Enuresis (Age 14) | 0.94 (0.78-1.12) | 0.48 |
| Intelligence | Soiling (Age 9) | 0.96 (0.85-1.08) | 0.48 |
| Intelligence | Soiling (Age 14) | 1.05 (0.91-1.20) | 0.53 |
| Intelligence | Constipation (Age 9) | 1.00 (0.91-1.09) | 0.96 |
| Intelligence | Constipation (Age 14) | 0.97 (0.87-1.08) | 0.60 |
| Anxiety | Daytime Urinary Incontinence (Age 9) | 1.02 (0.90-1.15) | 0.79 |
| Anxiety | Daytime Urinary Incontinence (Age 14) | 0.97 (0.82-1.14) | 0.69 |
| Anxiety | Enuresis (Age 9) | 0.93 (0.85-1.02) | 0.14 |
| Anxiety | Enuresis (Age 14) | 1.05 (0.88-1.26) | 0.57 |
| Anxiety | Soiling (Age 9) | 1.02 (0.90-1.14) | 0.80 |
| Anxiety | Soiling (Age 14) | 1.05 (0.92-1.21) | 0.47 |
| Anxiety | Constipation (Age 9) | 1.05 (0.96-1.15) | 0.27 |
| Anxiety | Constipation (Age 14) | 0.96 (0.87-1.07) | 0.47 |
| Depression | Daytime Urinary Incontinence (Age 9) | 1.22 (1.08-1.38) | 0.00 |
| Depression | Daytime Urinary Incontinence (Age 14) | 0.96 (0.82-1.14) | 0.67 |
| Depression | Enuresis (Age 9) | 1.00 (0.91-1.09) | 0.98 |
| Depression | Enuresis (Age 14) | 1.07 (0.89-1.28) | 0.49 |
| Depression | Soiling (Age 9) | 1.12 (0.99-1.26) | 0.07 |
| Depression | Soiling (Age 14) | 0.93 (0.81-1.06) | 0.27 |
| Depression | Constipation (Age 9) | 1.13 (1.03-1.24) | 0.01 |
| Depression | Constipation (Age 14) | 1.15 (1.03-1.28) | 0.01 |
| OCD | Daytime Urinary Incontinence (Age 9) | 0.89 (0.78-1.00) | 0.05 |
| OCD | Daytime Urinary Incontinence (Age 14) | 0.95 (0.80-1.12) | 0.55 |
| OCD | Enuresis (Age 9) | 0.97 (0.89-1.06) | 0.56 |
| OCD | Enuresis (Age 14) | 0.85 (0.71-1.02) | 0.08 |
| OCD | Soiling (Age 9) | 1.00 (0.89-1.13) | 0.97 |
| OCD | Soiling (Age 14) | 1.03 (0.90-1.19) | 0.66 |
| OCD | Constipation (Age 9) | 0.97 (0.89-1.06) | 0.53 |
| OCD | Constipation (Age 14) | 1.04 (0.94-1.16) | 0.42 |

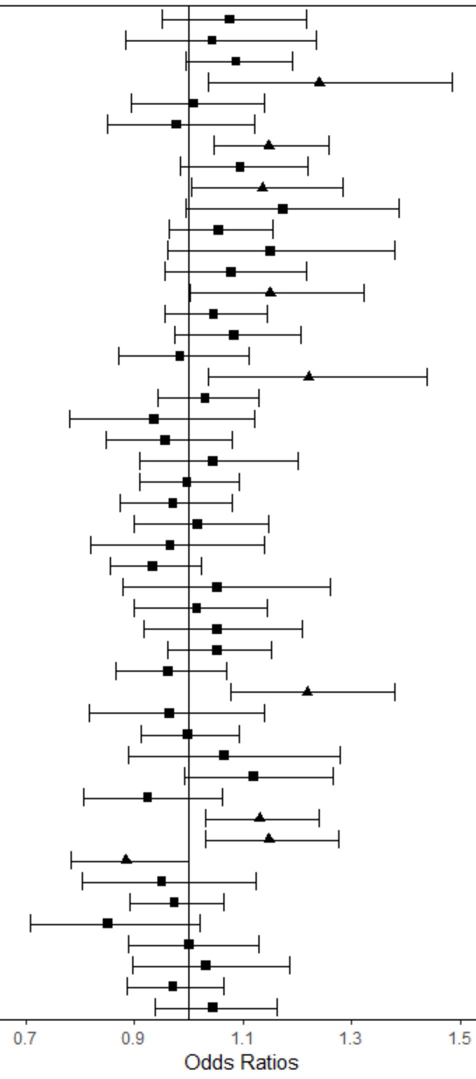

Supplementary Figure 2. Coefficients and 95% confidence intervals for the analysis of PRS Scores at the p < 0.4 SNP-inclusion significance level (S2)

| PRS | Incontinence Type | Odds Ratios (95% CI) | *p* |
|---|---|---|---|
| ADHD | Daytime Urinary Incontinence (Age 9) | 1.07 (0.95-1.21) | 0.28 |
| ADHD | Daytime Urinary Incontinence (Age 14) | 1.04 (0.88-1.23) | 0.66 |
| ADHD | Enuresis (Age 9) | 1.08 (0.99-1.18) | 0.09 |
| ADHD | Enuresis (Age 14) | 1.23 (1.03-1.47) | 0.03 |
| ADHD | Soiling (Age 9) | 1.01 (0.89-1.14) | 0.90 |
| ADHD | Soiling (Age 14) | 0.98 (0.86-1.13) | 0.81 |
| ADHD | Constipation (Age 9) | 1.15 (1.05-1.26) | 0.00 |
| ADHD | Constipation (Age 14) | 1.09 (0.98-1.21) | 0.11 |
| Autism | Daytime Urinary Incontinence (Age 9) | 1.14 (1.01-1.29) | 0.03 |
| Autism | Daytime Urinary Incontinence (Age 14) | 1.19 (1.01-1.40) | 0.04 |
| Autism | Enuresis (Age 9) | 1.06 (0.96-1.15) | 0.24 |
| Autism | Enuresis (Age 14) | 1.16 (0.97-1.39) | 0.11 |
| Autism | Soiling (Age 9) | 1.08 (0.96-1.22) | 0.22 |
| Autism | Soiling (Age 14) | 1.15 (1.00-1.32) | 0.04 |
| Autism | Constipation (Age 9) | 1.05 (0.96-1.14) | 0.34 |
| Autism | Constipation (Age 14) | 1.07 (0.96-1.19) | 0.21 |
| Intelligence | Daytime Urinary Incontinence (Age 9) | 0.99 (0.87-1.11) | 0.83 |
| Intelligence | Daytime Urinary Incontinence (Age 14) | 1.22 (1.03-1.43) | 0.02 |
| Intelligence | Enuresis (Age 9) | 1.03 (0.94-1.13) | 0.52 |
| Intelligence | Enuresis (Age 14) | 0.95 (0.79-1.14) | 0.56 |
| Intelligence | Soiling (Age 9) | 0.96 (0.85-1.09) | 0.55 |
| Intelligence | Soiling (Age 14) | 1.05 (0.91-1.20) | 0.51 |
| Intelligence | Constipation (Age 9) | 0.99 (0.91-1.09) | 0.90 |
| Intelligence | Constipation (Age 14) | 0.97 (0.87-1.08) | 0.59 |
| Anxiety | Daytime Urinary Incontinence (Age 9) | 1.03 (0.91-1.16) | 0.65 |
| Anxiety | Daytime Urinary Incontinence (Age 14) | 0.96 (0.81-1.13) | 0.62 |
| Anxiety | Enuresis (Age 9) | 0.93 (0.85-1.02) | 0.14 |
| Anxiety | Enuresis (Age 14) | 1.05 (0.88-1.26) | 0.57 |
| Anxiety | Soiling (Age 9) | 1.02 (0.90-1.15) | 0.77 |
| Anxiety | Soiling (Age 14) | 1.06 (0.92-1.21) | 0.45 |
| Anxiety | Constipation (Age 9) | 1.05 (0.96-1.15) | 0.26 |
| Anxiety | Constipation (Age 14) | 0.96 (0.86-1.06) | 0.40 |
| Depression | Daytime Urinary Incontinence (Age 9) | 1.21 (1.07-1.37) | 0.00 |
| Depression | Daytime Urinary Incontinence (Age 14) | 0.97 (0.82-1.14) | 0.69 |
| Depression | Enuresis (Age 9) | 1.00 (0.91-1.09) | 0.92 |
| Depression | Enuresis (Age 14) | 1.07 (0.89-1.28) | 0.47 |
| Depression | Soiling (Age 9) | 1.11 (0.99-1.26) | 0.08 |
| Depression | Soiling (Age 14) | 0.93 (0.81-1.06) | 0.27 |
| Depression | Constipation (Age 9) | 1.13 (1.03-1.23) | 0.01 |
| Depression | Constipation (Age 14) | 1.15 (1.03-1.28) | 0.01 |
| OCD | Daytime Urinary Incontinence (Age 9) | 0.89 (0.79-1.00) | 0.06 |
| OCD | Daytime Urinary Incontinence (Age 14) | 0.96 (0.81-1.13) | 0.59 |
| OCD | Enuresis (Age 9) | 0.98 (0.89-1.07) | 0.63 |
| OCD | Enuresis (Age 14) | 0.84 (0.70-1.01) | 0.06 |
| OCD | Soiling (Age 9) | 1.00 (0.89-1.13) | 1.00 |
| OCD | Soiling (Age 14) | 1.04 (0.90-1.19) | 0.60 |
| OCD | Constipation (Age 9) | 0.98 (0.89-1.07) | 0.61 |
| OCD | Constipation (Age 14) | 1.04 (0.93-1.15) | 0.48 |

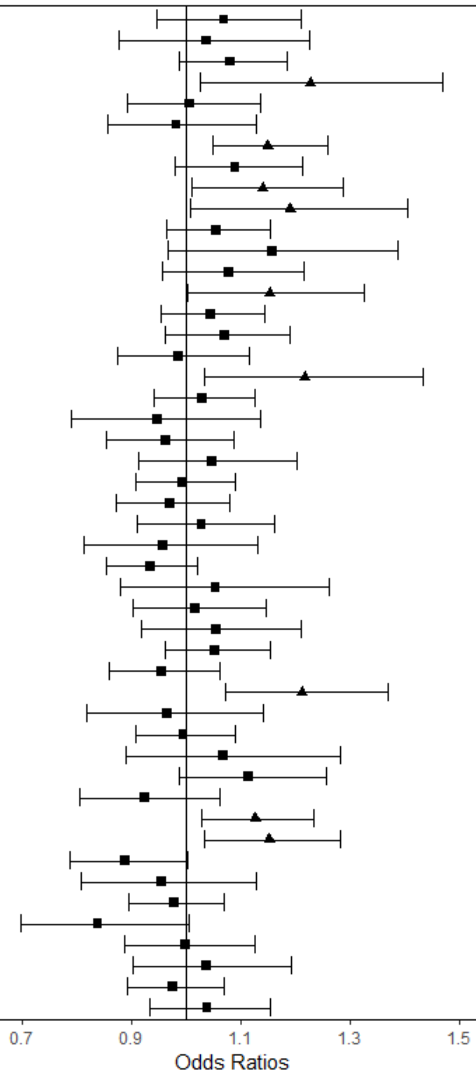

Supplementary Figure 3. Coefficients and 95% confidence intervals for the analysis of PRS Scores at the p < 0.3 SNP-inclusion significance level (S3)

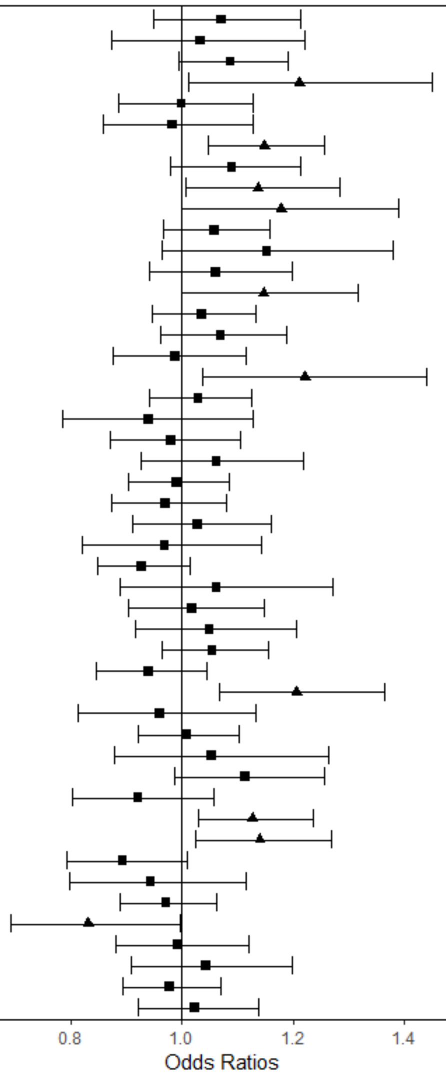


| PRS | Incontinence Type | Odds Ratios (95% CI) | *p* |
|---|---|---|---|
| ADHD | Daytime Urinary Incontinence (Age 9) | 1.07 (0.95-1.21) | 0.27 |
| ADHD | Daytime Urinary Incontinence (Age 14) | 1.03 (0.87-1.22) | 0.71 |
| ADHD | Enuresis (Age 9) | 1.09 (0.99-1.19) | 0.07 |
| ADHD | Enuresis (Age 14) | 1.21 (1.01-1.45) | 0.04 |
| ADHD | Soiling (Age 9) | 1.00 (0.89-1.13) | 0.98 |
| ADHD | Soiling (Age 14) | 0.98 (0.86-1.13) | 0.81 |
| ADHD | Constipation (Age 9) | 1.15 (1.05-1.26) | 0.00 |
| ADHD | Constipation (Age 14) | 1.09 (0.98-1.21) | 0.11 |
| Autism | Daytime Urinary Incontinence (Age 9) | 1.14 (1.01-1.28) | 0.04 |
| Autism | Daytime Urinary Incontinence (Age 14) | 1.18 (1.00-1.39) | 0.05 |
| Autism | Enuresis (Age 9) | 1.06 (0.97-1.16) | 0.22 |
| Autism | Enuresis (Age 14) | 1.15 (0.96-1.38) | 0.12 |
| Autism | Soiling (Age 9) | 1.06 (0.94-1.20) | 0.33 |
| Autism | Soiling (Age 14) | 1.15 (1.00-1.32) | 0.05 |
| Autism | Constipation (Age 9) | 1.04 (0.95-1.13) | 0.45 |
| Autism | Constipation (Age 14) | 1.07 (0.96-1.19) | 0.22 |
| Intelligence | Daytime Urinary Incontinence (Age 9) | 0.99 (0.88-1.12) | 0.85 |
| Intelligence | Daytime Urinary Incontinence (Age 14) | 1.22 (1.04-1.44) | 0.02 |
| Intelligence | Enuresis (Age 9) | 1.03 (0.94-1.13) | 0.53 |
| Intelligence | Enuresis (Age 14) | 0.94 (0.78-1.13) | 0.50 |
| Intelligence | Soiling (Age 9) | 0.98 (0.87-1.11) | 0.75 |
| Intelligence | Soiling (Age 14) | 1.06 (0.93-1.22) | 0.39 |
| Intelligence | Constipation (Age 9) | 0.99 (0.90-1.08) | 0.83 |
| Intelligence | Constipation (Age 14) | 0.97 (0.87-1.08) | 0.57 |
| Anxiety | Daytime Urinary Incontinence (Age 9) | 1.03 (0.91-1.16) | 0.66 |
| Anxiety | Daytime Urinary Incontinence (Age 14) | 0.97 (0.82-1.14) | 0.70 |
| Anxiety | Enuresis (Age 9) | 0.93 (0.85-1.01) | 0.10 |
| Anxiety | Enuresis (Age 14) | 1.06 (0.89-1.27) | 0.51 |
| Anxiety | Soiling (Age 9) | 1.02 (0.90-1.15) | 0.77 |
| Anxiety | Soiling (Age 14) | 1.05 (0.91-1.20) | 0.49 |
| Anxiety | Constipation (Age 9) | 1.05 (0.96-1.15) | 0.26 |
| Anxiety | Constipation (Age 14) | 0.94 (0.84-1.04) | 0.24 |
| Depression | Daytime Urinary Incontinence (Age 9) | 1.21 (1.07-1.36) | 0.00 |
| Depression | Daytime Urinary Incontinence (Age 14) | 0.96 (0.81-1.13) | 0.62 |
| Depression | Enuresis (Age 9) | 1.01 (0.92-1.10) | 0.88 |
| Depression | Enuresis (Age 14) | 1.05 (0.88-1.26) | 0.58 |
| Depression | Soiling (Age 9) | 1.11 (0.99-1.26) | 0.08 |
| Depression | Soiling (Age 14) | 0.92 (0.80-1.06) | 0.24 |
| Depression | Constipation (Age 9) | 1.13 (1.03-1.24) | 0.01 |
| Depression | Constipation (Age 14) | 1.14 (1.02-1.27) | 0.02 |
| OCD | Daytime Urinary Incontinence (Age 9) | 0.89 (0.79-1.01) | 0.07 |
| OCD | Daytime Urinary Incontinence (Age 14) | 0.94 (0.80-1.12) | 0.50 |
| OCD | Enuresis (Age 9) | 0.97 (0.89-1.06) | 0.52 |
| OCD | Enuresis (Age 14) | 0.83 (0.69-1.00) | 0.05 |
| OCD | Soiling (Age 9) | 0.99 (0.88-1.12) | 0.90 |
| OCD | Soiling (Age 14) | 1.04 (0.91-1.20) | 0.55 |
| OCD | Constipation (Age 9) | 0.98 (0.89-1.07) | 0.62 |
| OCD | Constipation (Age 14) | 1.02 (0.92-1.14) | 0.68 |

Supplementary Figure 4. Coefficients and 95% confidence intervals for the analysis of PRS Scores at the p < 0.2 SNP-inclusion significance level (S4)

| PRS | Incontinence Type | Odds Ratios (95% CI) | *p* |
|---|---|---|---|
| ADHD | Daytime Urinary Incontinence (Age 9) | 1.09 (0.96-1.23) | 0.19 |
| ADHD | Daytime Urinary Incontinence (Age 14) | 1.02 (0.86-1.21) | 0.82 |
| ADHD | Enuresis (Age 9) | 1.09 (0.99-1.19) | 0.07 |
| ADHD | Enuresis (Age 14) | 1.20 (1.01-1.44) | 0.04 |
| ADHD | Soiling (Age 9) | 1.00 (0.88-1.13) | 0.98 |
| ADHD | Soiling (Age 14) | 0.98 (0.85-1.12) | 0.73 |
| ADHD | Constipation (Age 9) | 1.15 (1.05-1.26) | 0.00 |
| ADHD | Constipation (Age 14) | 1.07 (0.97-1.19) | 0.19 |
| Autism | Daytime Urinary Incontinence (Age 9) | 1.13 (1.00-1.27) | 0.05 |
| Autism | Daytime Urinary Incontinence (Age 14) | 1.19 (1.01-1.41) | 0.04 |
| Autism | Enuresis (Age 9) | 1.06 (0.97-1.16) | 0.18 |
| Autism | Enuresis (Age 14) | 1.15 (0.96-1.38) | 0.13 |
| Autism | Soiling (Age 9) | 1.03 (0.92-1.17) | 0.59 |
| Autism | Soiling (Age 14) | 1.14 (1.00-1.31) | 0.06 |
| Autism | Constipation (Age 9) | 1.03 (0.94-1.13) | 0.47 |
| Autism | Constipation (Age 14) | 1.07 (0.96-1.19) | 0.21 |
| Intelligence | Daytime Urinary Incontinence (Age 9) | 0.98 (0.87-1.11) | 0.78 |
| Intelligence | Daytime Urinary Incontinence (Age 14) | 1.18 (1.00-1.39) | 0.05 |
| Intelligence | Enuresis (Age 9) | 1.03 (0.94-1.13) | 0.52 |
| Intelligence | Enuresis (Age 14) | 0.94 (0.78-1.12) | 0.48 |
| Intelligence | Soiling (Age 9) | 0.98 (0.87-1.11) | 0.77 |
| Intelligence | Soiling (Age 14) | 1.07 (0.93-1.23) | 0.32 |
| Intelligence | Constipation (Age 9) | 1.00 (0.91-1.09) | 0.97 |
| Intelligence | Constipation (Age 14) | 0.98 (0.88-1.09) | 0.73 |
| Anxiety | Daytime Urinary Incontinence (Age 9) | 1.02 (0.90-1.15) | 0.77 |
| Anxiety | Daytime Urinary Incontinence (Age 14) | 0.98 (0.83-1.16) | 0.83 |
| Anxiety | Enuresis (Age 9) | 0.91 (0.83-1.00) | 0.04 |
| Anxiety | Enuresis (Age 14) | 1.06 (0.88-1.26) | 0.55 |
| Anxiety | Soiling (Age 9) | 1.01 (0.90-1.14) | 0.82 |
| Anxiety | Soiling (Age 14) | 1.08 (0.94-1.24) | 0.30 |
| Anxiety | Constipation (Age 9) | 1.08 (0.98-1.18) | 0.11 |
| Anxiety | Constipation (Age 14) | 0.95 (0.86-1.06) | 0.38 |
| Depression | Daytime Urinary Incontinence (Age 9) | 1.21 (1.07-1.36) | 0.00 |
| Depression | Daytime Urinary Incontinence (Age 14) | 0.94 (0.80-1.11) | 0.48 |
| Depression | Enuresis (Age 9) | 1.01 (0.93-1.11) | 0.77 |
| Depression | Enuresis (Age 14) | 1.04 (0.87-1.25) | 0.67 |
| Depression | Soiling (Age 9) | 1.10 (0.97-1.24) | 0.12 |
| Depression | Soiling (Age 14) | 0.92 (0.80-1.05) | 0.23 |
| Depression | Constipation (Age 9) | 1.12 (1.02-1.23) | 0.01 |
| Depression | Constipation (Age 14) | 1.12 (1.01-1.25) | 0.03 |
| OCD | Daytime Urinary Incontinence (Age 9) | 0.92 (0.82-1.04) | 0.19 |
| OCD | Daytime Urinary Incontinence (Age 14) | 0.94 (0.79-1.11) | 0.45 |
| OCD | Enuresis (Age 9) | 0.99 (0.90-1.08) | 0.77 |
| OCD | Enuresis (Age 14) | 0.86 (0.72-1.03) | 0.11 |
| OCD | Soiling (Age 9) | 0.99 (0.88-1.11) | 0.85 |
| OCD | Soiling (Age 14) | 1.02 (0.89-1.17) | 0.75 |
| OCD | Constipation (Age 9) | 0.98 (0.89-1.07) | 0.65 |
| OCD | Constipation (Age 14) | 1.01 (0.91-1.12) | 0.89 |

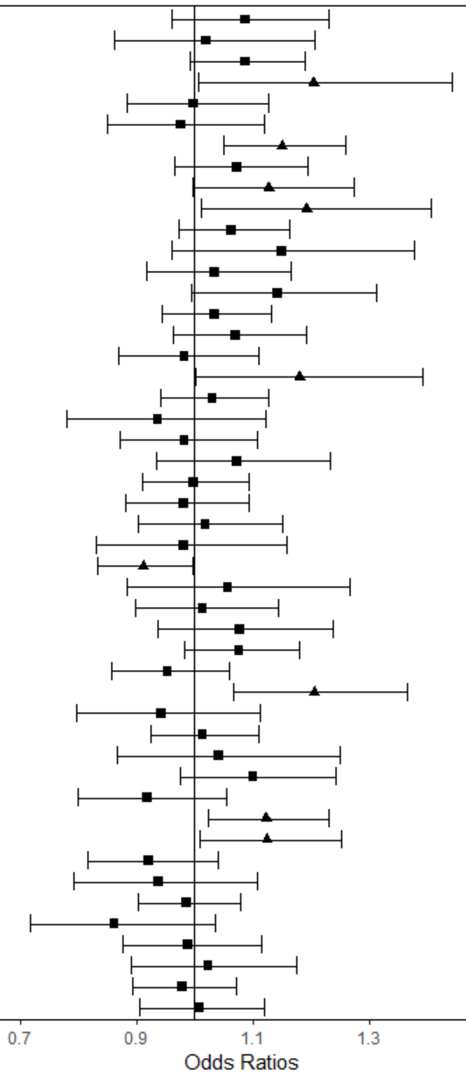

Supplementary Figure 5. Coefficients and 95% confidence intervals for the analysis of PRS Scores at the p < 0.1 SNP-inclusion significance level (S5)

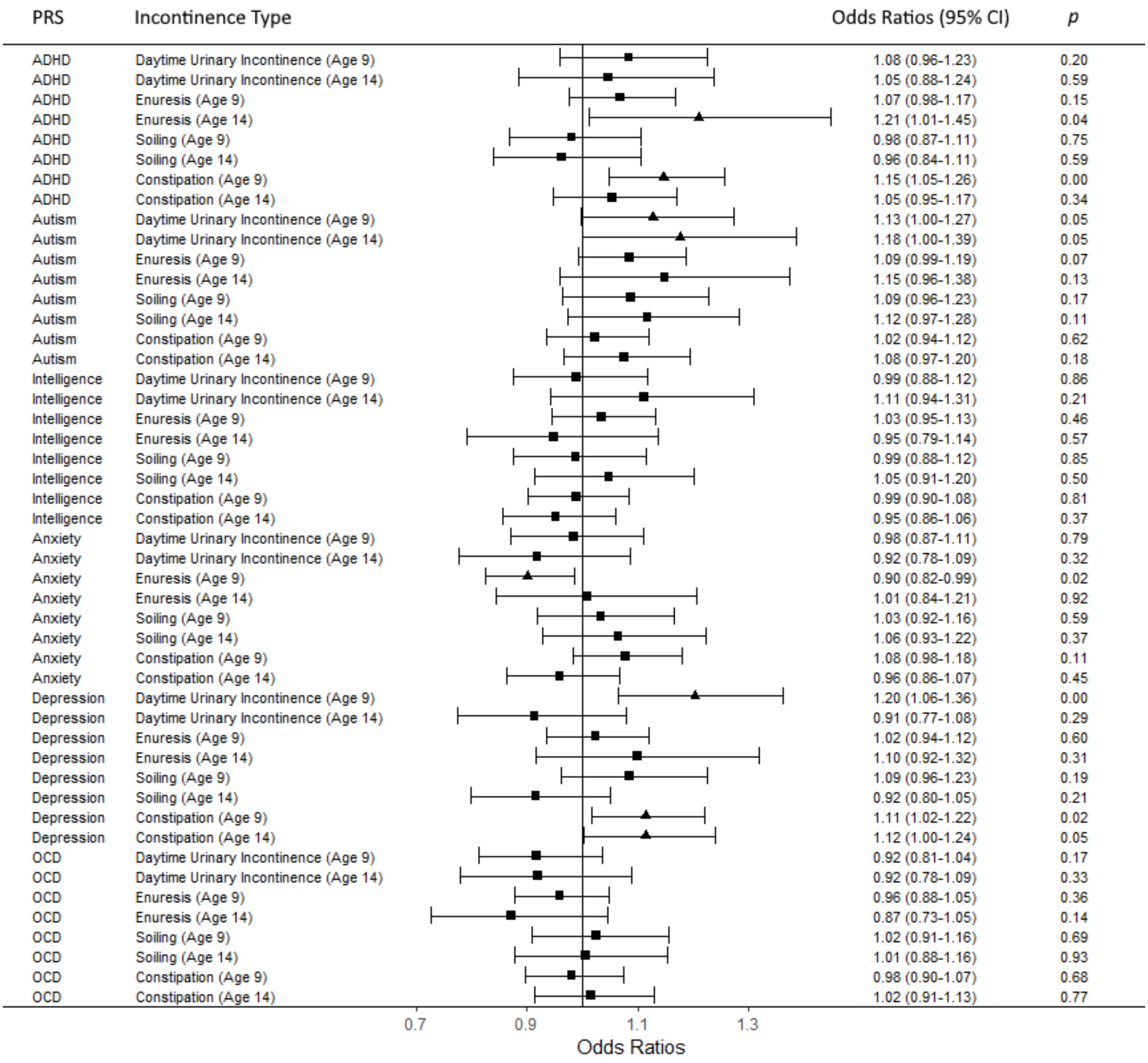

Supplementary Figure 6. Coefficients and 95% confidence intervals for the analysis of PRS Scores at the $p < 0.05$ SNP-inclusion significance level (S6)

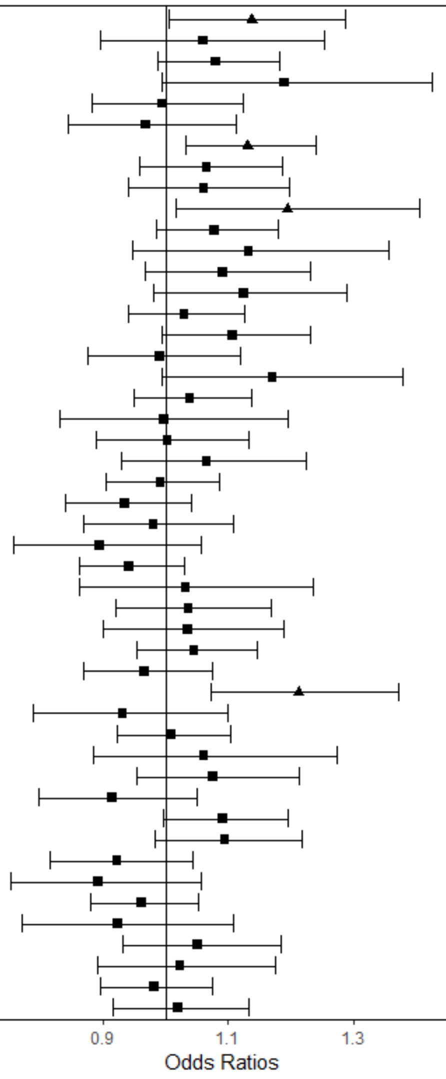


| PRS | Incontinence Type | Odds Ratios (95% CI) | *p* |
|---|---|---|---|
| ADHD | Daytime Urinary Incontinence (Age 9) | 1.14 (1.01-1.29) | 0.04 |
| ADHD | Daytime Urinary Incontinence (Age 14) | 1.06 (0.90-1.25) | 0.50 |
| ADHD | Enuresis (Age 9) | 1.08 (0.99-1.18) | 0.10 |
| ADHD | Enuresis (Age 14) | 1.19 (0.99-1.43) | 0.06 |
| ADHD | Soiling (Age 9) | 1.00 (0.88-1.12) | 0.94 |
| ADHD | Soiling (Age 14) | 0.97 (0.84-1.11) | 0.65 |
| ADHD | Constipation (Age 9) | 1.13 (1.03-1.24) | 0.01 |
| ADHD | Constipation (Age 14) | 1.07 (0.96-1.19) | 0.24 |
| Autism | Daytime Urinary Incontinence (Age 9) | 1.06 (0.94-1.20) | 0.34 |
| Autism | Daytime Urinary Incontinence (Age 14) | 1.19 (1.02-1.41) | 0.03 |
| Autism | Enuresis (Age 9) | 1.08 (0.98-1.18) | 0.11 |
| Autism | Enuresis (Age 14) | 1.13 (0.95-1.36) | 0.17 |
| Autism | Soiling (Age 9) | 1.09 (0.97-1.23) | 0.16 |
| Autism | Soiling (Age 14) | 1.12 (0.98-1.29) | 0.09 |
| Autism | Constipation (Age 9) | 1.03 (0.94-1.13) | 0.53 |
| Autism | Constipation (Age 14) | 1.11 (0.99-1.23) | 0.06 |
| Intelligence | Daytime Urinary Incontinence (Age 9) | 0.99 (0.88-1.12) | 0.87 |
| Intelligence | Daytime Urinary Incontinence (Age 14) | 1.17 (0.99-1.38) | 0.06 |
| Intelligence | Enuresis (Age 9) | 1.04 (0.95-1.14) | 0.41 |
| Intelligence | Enuresis (Age 14) | 1.00 (0.83-1.20) | 0.97 |
| Intelligence | Soiling (Age 9) | 1.00 (0.89-1.13) | 0.96 |
| Intelligence | Soiling (Age 14) | 1.07 (0.93-1.22) | 0.37 |
| Intelligence | Constipation (Age 9) | 0.99 (0.90-1.09) | 0.85 |
| Intelligence | Constipation (Age 14) | 0.93 (0.84-1.04) | 0.22 |
| Anxiety | Daytime Urinary Incontinence (Age 9) | 0.98 (0.87-1.11) | 0.75 |
| Anxiety | Daytime Urinary Incontinence (Age 14) | 0.89 (0.76-1.06) | 0.19 |
| Anxiety | Enuresis (Age 9) | 0.94 (0.86-1.03) | 0.19 |
| Anxiety | Enuresis (Age 14) | 1.03 (0.86-1.23) | 0.73 |
| Anxiety | Soiling (Age 9) | 1.04 (0.92-1.17) | 0.56 |
| Anxiety | Soiling (Age 14) | 1.03 (0.90-1.19) | 0.63 |
| Anxiety | Constipation (Age 9) | 1.05 (0.95-1.14) | 0.34 |
| Anxiety | Constipation (Age 14) | 0.97 (0.87-1.07) | 0.52 |
| Depression | Daytime Urinary Incontinence (Age 9) | 1.21 (1.07-1.37) | 0.00 |
| Depression | Daytime Urinary Incontinence (Age 14) | 0.93 (0.79-1.10) | 0.40 |
| Depression | Enuresis (Age 9) | 1.01 (0.92-1.10) | 0.86 |
| Depression | Enuresis (Age 14) | 1.06 (0.88-1.27) | 0.52 |
| Depression | Soiling (Age 9) | 1.08 (0.95-1.21) | 0.24 |
| Depression | Soiling (Age 14) | 0.91 (0.80-1.05) | 0.20 |
| Depression | Constipation (Age 9) | 1.09 (1.00-1.20) | 0.06 |
| Depression | Constipation (Age 14) | 1.09 (0.98-1.22) | 0.10 |
| OCD | Daytime Urinary Incontinence (Age 9) | 0.92 (0.82-1.04) | 0.20 |
| OCD | Daytime Urinary Incontinence (Age 14) | 0.89 (0.75-1.06) | 0.18 |
| OCD | Enuresis (Age 9) | 0.96 (0.88-1.05) | 0.40 |
| OCD | Enuresis (Age 14) | 0.92 (0.77-1.11) | 0.39 |
| OCD | Soiling (Age 9) | 1.05 (0.93-1.18) | 0.42 |
| OCD | Soiling (Age 14) | 1.02 (0.89-1.17) | 0.75 |
| OCD | Constipation (Age 9) | 0.98 (0.90-1.07) | 0.68 |
| OCD | Constipation (Age 14) | 1.02 (0.92-1.13) | 0.73 |

Supplementary Figure 7. Coefficients and 95% confidence intervals for the analysis of PRS Scores at the p < 0.01 SNP-inclusion significance level (S7)

| PRS | Incontinence Type | Odds Ratios (95% CI) | *p* |
|---|---|---|---|
| ADHD | Daytime Urinary Incontinence (Age 9) | 1.13 (1.00-1.28) | 0.05 |
| ADHD | Daytime Urinary Incontinence (Age 14) | 0.98 (0.83-1.16) | 0.81 |
| ADHD | Enuresis (Age 9) | 1.04 (0.95-1.13) | 0.44 |
| ADHD | Enuresis (Age 14) | 1.12 (0.94-1.34) | 0.21 |
| ADHD | Soiling (Age 9) | 1.04 (0.93-1.18) | 0.48 |
| ADHD | Soiling (Age 14) | 1.00 (0.87-1.15) | 0.96 |
| ADHD | Constipation (Age 9) | 1.08 (0.98-1.18) | 0.11 |
| ADHD | Constipation (Age 14) | 1.06 (0.95-1.18) | 0.31 |
| Autism | Daytime Urinary Incontinence (Age 9) | 1.05 (0.93-1.18) | 0.45 |
| Autism | Daytime Urinary Incontinence (Age 14) | 1.13 (0.96-1.33) | 0.13 |
| Autism | Enuresis (Age 9) | 1.03 (0.94-1.13) | 0.47 |
| Autism | Enuresis (Age 14) | 1.15 (0.96-1.38) | 0.12 |
| Autism | Soiling (Age 9) | 0.99 (0.88-1.12) | 0.90 |
| Autism | Soiling (Age 14) | 1.01 (0.88-1.16) | 0.85 |
| Autism | Constipation (Age 9) | 1.04 (0.95-1.13) | 0.44 |
| Autism | Constipation (Age 14) | 1.11 (1.00-1.23) | 0.06 |
| Intelligence | Daytime Urinary Incontinence (Age 9) | 1.04 (0.92-1.17) | 0.56 |
| Intelligence | Daytime Urinary Incontinence (Age 14) | 1.20 (1.01-1.41) | 0.03 |
| Intelligence | Enuresis (Age 9) | 1.09 (1.00-1.19) | 0.06 |
| Intelligence | Enuresis (Age 14) | 1.04 (0.87-1.25) | 0.68 |
| Intelligence | Soiling (Age 9) | 0.98 (0.87-1.11) | 0.73 |
| Intelligence | Soiling (Age 14) | 1.20 (1.04-1.38) | 0.01 |
| Intelligence | Constipation (Age 9) | 0.97 (0.89-1.07) | 0.56 |
| Intelligence | Constipation (Age 14) | 0.94 (0.85-1.05) | 0.29 |
| Anxiety | Daytime Urinary Incontinence (Age 9) | 1.03 (0.91-1.16) | 0.63 |
| Anxiety | Daytime Urinary Incontinence (Age 14) | 0.88 (0.74-1.04) | 0.12 |
| Anxiety | Enuresis (Age 9) | 0.94 (0.86-1.03) | 0.16 |
| Anxiety | Enuresis (Age 14) | 1.04 (0.87-1.25) | 0.64 |
| Anxiety | Soiling (Age 9) | 1.04 (0.92-1.17) | 0.52 |
| Anxiety | Soiling (Age 14) | 1.02 (0.89-1.17) | 0.81 |
| Anxiety | Constipation (Age 9) | 1.03 (0.94-1.12) | 0.58 |
| Anxiety | Constipation (Age 14) | 1.03 (0.93-1.15) | 0.59 |
| Depression | Daytime Urinary Incontinence (Age 9) | 1.23 (1.09-1.39) | 0.00 |
| Depression | Daytime Urinary Incontinence (Age 14) | 0.95 (0.80-1.12) | 0.53 |
| Depression | Enuresis (Age 9) | 1.01 (0.92-1.10) | 0.88 |
| Depression | Enuresis (Age 14) | 1.12 (0.93-1.35) | 0.23 |
| Depression | Soiling (Age 9) | 1.11 (0.98-1.25) | 0.10 |
| Depression | Soiling (Age 14) | 0.92 (0.80-1.06) | 0.27 |
| Depression | Constipation (Age 9) | 1.12 (1.02-1.23) | 0.02 |
| Depression | Constipation (Age 14) | 1.08 (0.97-1.20) | 0.17 |
| OCD | Daytime Urinary Incontinence (Age 9) | 0.99 (0.88-1.12) | 0.90 |
| OCD | Daytime Urinary Incontinence (Age 14) | 0.93 (0.78-1.10) | 0.39 |
| OCD | Enuresis (Age 9) | 0.93 (0.85-1.02) | 0.11 |
| OCD | Enuresis (Age 14) | 0.83 (0.69-0.99) | 0.04 |
| OCD | Soiling (Age 9) | 1.15 (1.02-1.29) | 0.03 |
| OCD | Soiling (Age 14) | 0.94 (0.82-1.08) | 0.42 |
| OCD | Constipation (Age 9) | 0.95 (0.87-1.05) | 0.32 |
| OCD | Constipation (Age 14) | 1.08 (0.97-1.20) | 0.16 |

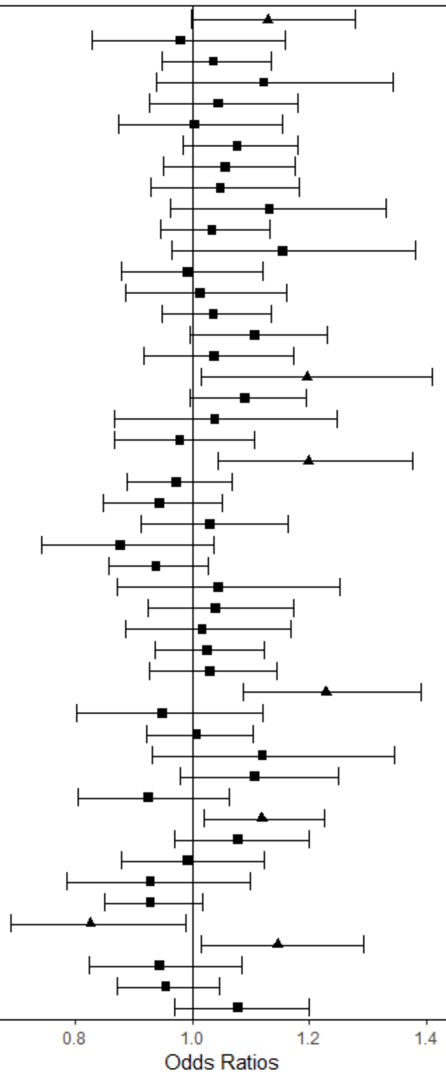

Supplementary Figure 8. Coefficients and 95% confidence intervals for the analysis of PRS Scores at the p < 0.001 SNP-inclusion significance level (S8)

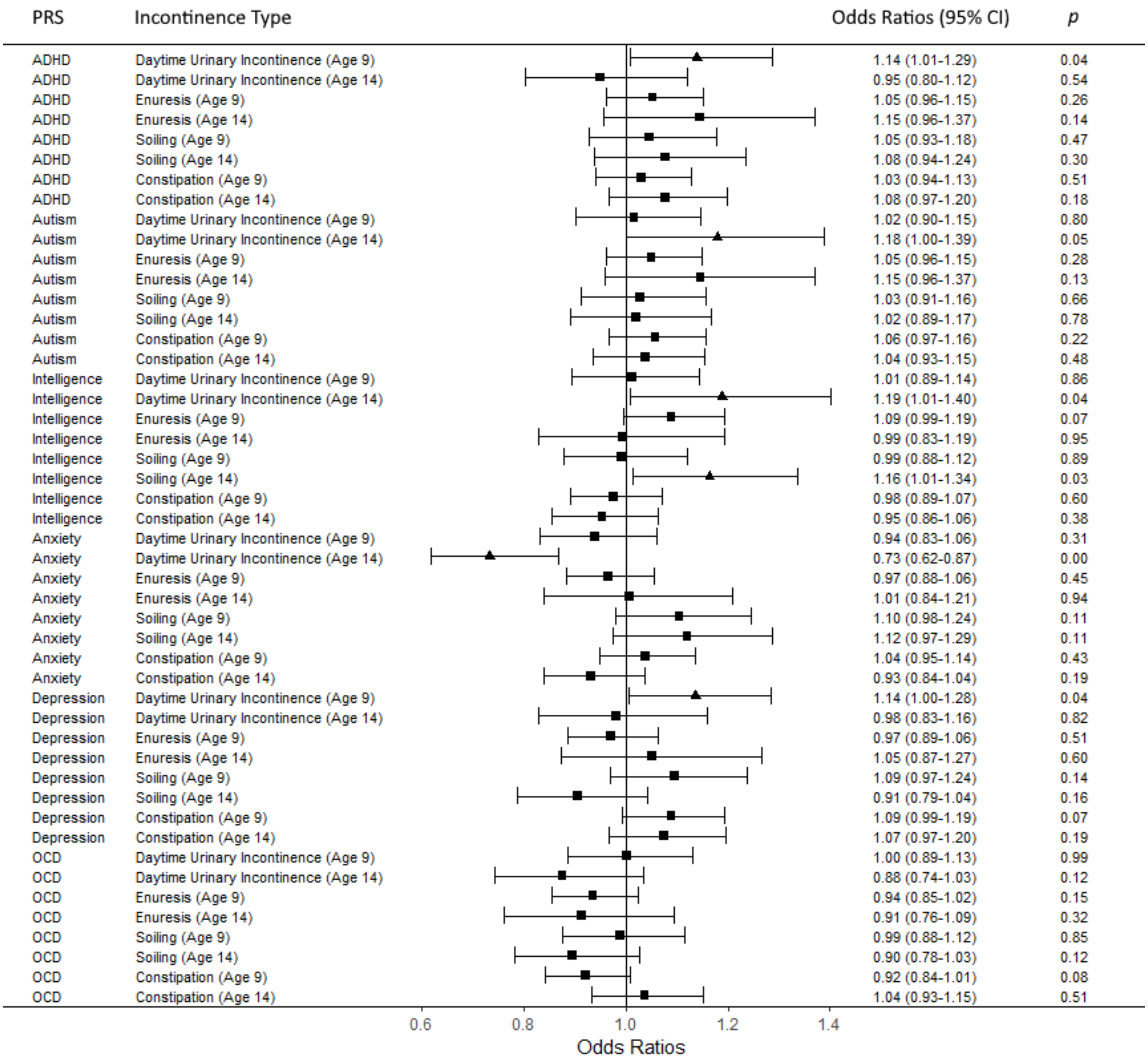

Supplementary Figure 9. Coefficients and 95% confidence intervals for the analysis of PRS Scores at the p < 0.0001 SNP-inclusion significance level (S9)

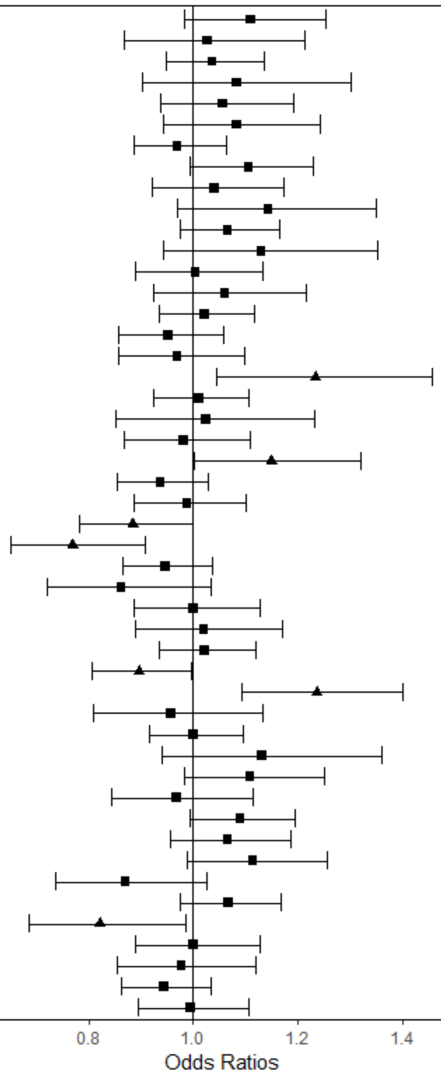


| PRS | Incontinence Type | Odds Ratios (95% CI) | *p* |
|---|---|---|---|
| ADHD | Daytime Urinary Incontinence (Age 9) | 1.11 (0.98-1.25) | 0.09 |
| ADHD | Daytime Urinary Incontinence (Age 14) | 1.03 (0.87-1.21) | 0.76 |
| ADHD | Enuresis (Age 9) | 1.04 (0.95-1.13) | 0.43 |
| ADHD | Enuresis (Age 14) | 1.08 (0.90-1.30) | 0.38 |
| ADHD | Soiling (Age 9) | 1.06 (0.94-1.19) | 0.37 |
| ADHD | Soiling (Age 14) | 1.08 (0.94-1.24) | 0.26 |
| ADHD | Constipation (Age 9) | 0.97 (0.89-1.06) | 0.51 |
| ADHD | Constipation (Age 14) | 1.11 (0.99-1.23) | 0.07 |
| Autism | Daytime Urinary Incontinence (Age 9) | 1.04 (0.92-1.17) | 0.53 |
| Autism | Daytime Urinary Incontinence (Age 14) | 1.14 (0.97-1.35) | 0.11 |
| Autism | Enuresis (Age 9) | 1.07 (0.97-1.17) | 0.17 |
| Autism | Enuresis (Age 14) | 1.13 (0.94-1.35) | 0.18 |
| Autism | Soiling (Age 9) | 1.00 (0.89-1.13) | 0.95 |
| Autism | Soiling (Age 14) | 1.06 (0.93-1.22) | 0.40 |
| Autism | Constipation (Age 9) | 1.02 (0.93-1.12) | 0.64 |
| Autism | Constipation (Age 14) | 0.95 (0.86-1.06) | 0.36 |
| Intelligence | Daytime Urinary Incontinence (Age 9) | 0.97 (0.86-1.10) | 0.63 |
| Intelligence | Daytime Urinary Incontinence (Age 14) | 1.23 (1.04-1.46) | 0.01 |
| Intelligence | Enuresis (Age 9) | 1.01 (0.92-1.11) | 0.81 |
| Intelligence | Enuresis (Age 14) | 1.02 (0.85-1.23) | 0.80 |
| Intelligence | Soiling (Age 9) | 0.98 (0.87-1.11) | 0.76 |
| Intelligence | Soiling (Age 14) | 1.15 (1.00-1.32) | 0.05 |
| Intelligence | Constipation (Age 9) | 0.94 (0.86-1.03) | 0.17 |
| Intelligence | Constipation (Age 14) | 0.99 (0.89-1.10) | 0.83 |
| Anxiety | Daytime Urinary Incontinence (Age 9) | 0.88 (0.78-1.00) | 0.05 |
| Anxiety | Daytime Urinary Incontinence (Age 14) | 0.77 (0.65-0.91) | 0.00 |
| Anxiety | Enuresis (Age 9) | 0.95 (0.87-1.04) | 0.24 |
| Anxiety | Enuresis (Age 14) | 0.86 (0.72-1.03) | 0.11 |
| Anxiety | Soiling (Age 9) | 1.00 (0.89-1.13) | 1.00 |
| Anxiety | Soiling (Age 14) | 1.02 (0.89-1.17) | 0.77 |
| Anxiety | Constipation (Age 9) | 1.02 (0.93-1.12) | 0.63 |
| Anxiety | Constipation (Age 14) | 0.90 (0.81-1.00) | 0.04 |
| Depression | Daytime Urinary Incontinence (Age 9) | 1.24 (1.09-1.40) | 0.00 |
| Depression | Daytime Urinary Incontinence (Age 14) | 0.96 (0.81-1.13) | 0.62 |
| Depression | Enuresis (Age 9) | 1.00 (0.91-1.10) | 0.98 |
| Depression | Enuresis (Age 14) | 1.13 (0.94-1.36) | 0.19 |
| Depression | Soiling (Age 9) | 1.11 (0.98-1.25) | 0.10 |
| Depression | Soiling (Age 14) | 0.97 (0.84-1.11) | 0.66 |
| Depression | Constipation (Age 9) | 1.09 (0.99-1.19) | 0.07 |
| Depression | Constipation (Age 14) | 1.07 (0.96-1.19) | 0.25 |
| OCD | Daytime Urinary Incontinence (Age 9) | 1.11 (0.99-1.26) | 0.08 |
| OCD | Daytime Urinary Incontinence (Age 14) | 0.87 (0.74-1.03) | 0.10 |
| OCD | Enuresis (Age 9) | 1.07 (0.98-1.17) | 0.15 |
| OCD | Enuresis (Age 14) | 0.82 (0.69-0.99) | 0.03 |
| OCD | Soiling (Age 9) | 1.00 (0.89-1.13) | 0.99 |
| OCD | Soiling (Age 14) | 0.98 (0.85-1.12) | 0.74 |
| OCD | Constipation (Age 9) | 0.94 (0.86-1.03) | 0.21 |
| OCD | Constipation (Age 14) | 0.99 (0.90-1.11) | 0.92 |

Supplementary Figure 10. Coefficients and 95% confidence intervals for the analysis of PRS Scores at the p < 0.00001 SNP-inclusion significance level (S10)

| PRS | Incontinence Type | Odds Ratios (95% CI) | *p* |
|---|---|---|---|
| ADHD | Daytime Urinary Incontinence (Age 9) | 1.18 (1.05-1.34) | 0.01 |
| ADHD | Daytime Urinary Incontinence (Age 14) | 1.01 (0.85-1.19) | 0.93 |
| ADHD | Enuresis (Age 9) | 1.03 (0.94-1.13) | 0.49 |
| ADHD | Enuresis (Age 14) | 1.11 (0.92-1.33) | 0.27 |
| ADHD | Soiling (Age 9) | 1.10 (0.98-1.24) | 0.12 |
| ADHD | Soiling (Age 14) | 1.09 (0.94-1.25) | 0.25 |
| ADHD | Constipation (Age 9) | 0.98 (0.90-1.08) | 0.69 |
| ADHD | Constipation (Age 14) | 1.09 (0.98-1.22) | 0.11 |
| Autism | Daytime Urinary Incontinence (Age 9) | 0.97 (0.86-1.09) | 0.61 |
| Autism | Daytime Urinary Incontinence (Age 14) | 1.11 (0.94-1.31) | 0.20 |
| Autism | Enuresis (Age 9) | 1.06 (0.97-1.16) | 0.22 |
| Autism | Enuresis (Age 14) | 1.17 (0.98-1.40) | 0.09 |
| Autism | Soiling (Age 9) | 0.99 (0.88-1.12) | 0.88 |
| Autism | Soiling (Age 14) | 0.97 (0.85-1.11) | 0.68 |
| Autism | Constipation (Age 9) | 1.01 (0.93-1.11) | 0.76 |
| Autism | Constipation (Age 14) | 1.05 (0.95-1.17) | 0.33 |
| Intelligence | Daytime Urinary Incontinence (Age 9) | 0.99 (0.87-1.12) | 0.84 |
| Intelligence | Daytime Urinary Incontinence (Age 14) | 1.15 (0.97-1.36) | 0.10 |
| Intelligence | Enuresis (Age 9) | 0.99 (0.91-1.09) | 0.91 |
| Intelligence | Enuresis (Age 14) | 1.01 (0.84-1.22) | 0.89 |
| Intelligence | Soiling (Age 9) | 0.99 (0.88-1.12) | 0.92 |
| Intelligence | Soiling (Age 14) | 1.12 (0.97-1.28) | 0.12 |
| Intelligence | Constipation (Age 9) | 1.01 (0.93-1.11) | 0.77 |
| Intelligence | Constipation (Age 14) | 1.04 (0.93-1.16) | 0.46 |
| Anxiety | Daytime Urinary Incontinence (Age 9) | 1.04 (0.92-1.18) | 0.52 |
| Anxiety | Daytime Urinary Incontinence (Age 14) | 0.95 (0.80-1.12) | 0.52 |
| Anxiety | Enuresis (Age 9) | 0.95 (0.87-1.04) | 0.27 |
| Anxiety | Enuresis (Age 14) | 1.15 (0.96-1.38) | 0.13 |
| Anxiety | Soiling (Age 9) | 1.09 (0.96-1.22) | 0.18 |
| Anxiety | Soiling (Age 14) | 0.98 (0.86-1.13) | 0.82 |
| Anxiety | Constipation (Age 9) | 0.95 (0.87-1.04) | 0.31 |
| Anxiety | Constipation (Age 14) | 0.95 (0.85-1.06) | 0.36 |
| Depression | Daytime Urinary Incontinence (Age 9) | 1.31 (1.16-1.49) | 0.00 |
| Depression | Daytime Urinary Incontinence (Age 14) | 1.01 (0.85-1.19) | 0.95 |
| Depression | Enuresis (Age 9) | 1.02 (0.93-1.12) | 0.62 |
| Depression | Enuresis (Age 14) | 1.18 (0.98-1.43) | 0.07 |
| Depression | Soiling (Age 9) | 1.11 (0.98-1.25) | 0.10 |
| Depression | Soiling (Age 14) | 0.99 (0.86-1.14) | 0.90 |
| Depression | Constipation (Age 9) | 1.08 (0.98-1.18) | 0.11 |
| Depression | Constipation (Age 14) | 1.04 (0.93-1.15) | 0.53 |
| OCD | Daytime Urinary Incontinence (Age 9) | 1.00 (0.89-1.13) | 0.96 |
| OCD | Daytime Urinary Incontinence (Age 14) | 1.02 (0.86-1.21) | 0.81 |
| OCD | Enuresis (Age 9) | 1.00 (0.91-1.09) | 1.00 |
| OCD | Enuresis (Age 14) | 1.02 (0.86-1.23) | 0.79 |
| OCD | Soiling (Age 9) | 0.94 (0.84-1.07) | 0.35 |
| OCD | Soiling (Age 14) | 1.02 (0.89-1.17) | 0.78 |
| OCD | Constipation (Age 9) | 1.01 (0.93-1.11) | 0.75 |
| OCD | Constipation (Age 14) | 0.95 (0.85-1.05) | 0.32 |

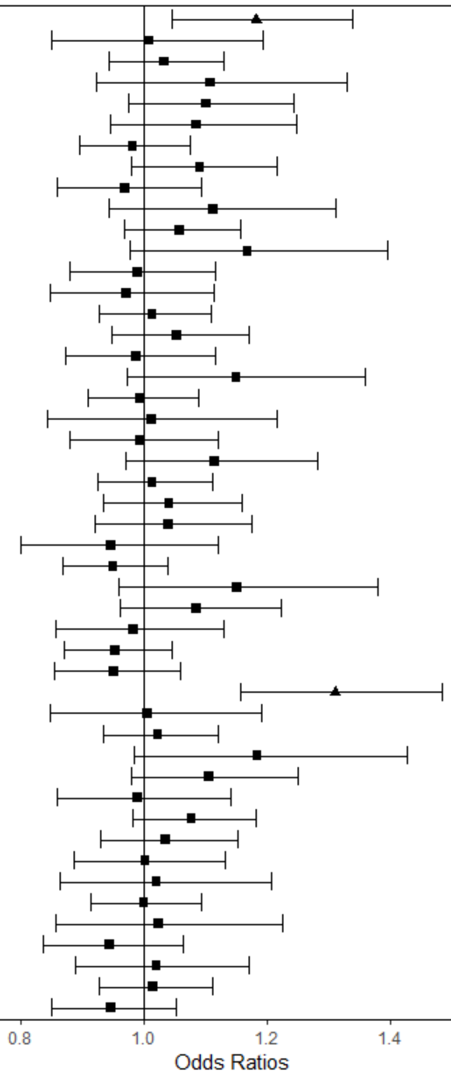

Supplementary Figure 11. Coefficients and 95% confidence intervals for the analysis of PRS Scores at the p < 0.000001 SNP-inclusion significance level (S11)

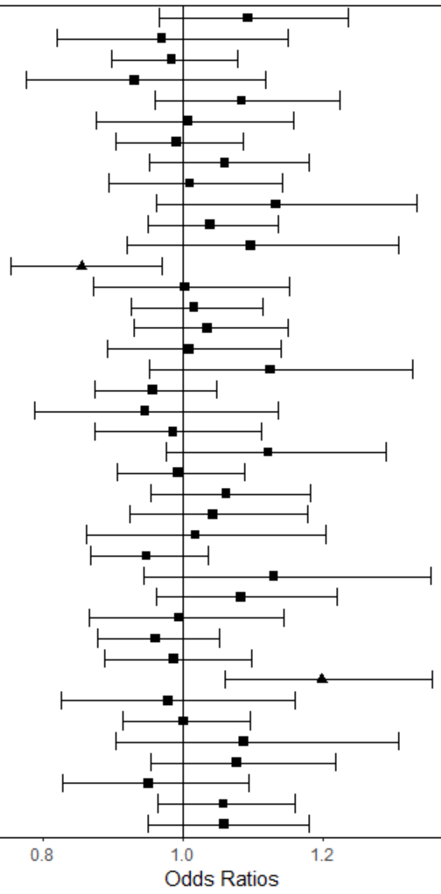


| PRS | Incontinence Type | Odds Ratios (95% CI) | *p* |
|---|---|---|---|
| ADHD | Daytime Urinary Incontinence (Age 9) | 1.09 (0.97-1.24) | 0.15 |
| ADHD | Daytime Urinary Incontinence (Age 14) | 0.97 (0.82-1.15) | 0.73 |
| ADHD | Enuresis (Age 9) | 0.98 (0.90-1.08) | 0.74 |
| ADHD | Enuresis (Age 14) | 0.93 (0.78-1.12) | 0.45 |
| ADHD | Soiling (Age 9) | 1.08 (0.96-1.22) | 0.19 |
| ADHD | Soiling (Age 14) | 1.01 (0.88-1.16) | 0.91 |
| ADHD | Constipation (Age 9) | 0.99 (0.91-1.09) | 0.86 |
| ADHD | Constipation (Age 14) | 1.06 (0.95-1.18) | 0.28 |
| Autism | Daytime Urinary Incontinence (Age 9) | 1.01 (0.89-1.14) | 0.87 |
| Autism | Daytime Urinary Incontinence (Age 14) | 1.13 (0.96-1.33) | 0.13 |
| Autism | Enuresis (Age 9) | 1.04 (0.95-1.14) | 0.39 |
| Autism | Enuresis (Age 14) | 1.10 (0.92-1.31) | 0.30 |
| Autism | Soiling (Age 9) | 0.86 (0.76-0.97) | 0.01 |
| Autism | Soiling (Age 14) | 1.00 (0.87-1.15) | 0.97 |
| Autism | Constipation (Age 9) | 1.02 (0.93-1.11) | 0.73 |
| Autism | Constipation (Age 14) | 1.03 (0.93-1.15) | 0.53 |
| Intelligence | Daytime Urinary Incontinence (Age 9) | 1.01 (0.89-1.14) | 0.88 |
| Intelligence | Daytime Urinary Incontinence (Age 14) | 1.12 (0.95-1.33) | 0.17 |
| Intelligence | Enuresis (Age 9) | 0.96 (0.88-1.05) | 0.34 |
| Intelligence | Enuresis (Age 14) | 0.95 (0.79-1.14) | 0.56 |
| Intelligence | Soiling (Age 9) | 0.99 (0.87-1.11) | 0.82 |
| Intelligence | Soiling (Age 14) | 1.12 (0.98-1.29) | 0.11 |
| Intelligence | Constipation (Age 9) | 0.99 (0.91-1.09) | 0.89 |
| Intelligence | Constipation (Age 14) | 1.06 (0.95-1.18) | 0.27 |
| Anxiety | Daytime Urinary Incontinence (Age 9) | 1.04 (0.93-1.18) | 0.49 |
| Anxiety | Daytime Urinary Incontinence (Age 14) | 1.02 (0.86-1.20) | 0.83 |
| Anxiety | Enuresis (Age 9) | 0.95 (0.87-1.04) | 0.25 |
| Anxiety | Enuresis (Age 14) | 1.13 (0.94-1.35) | 0.18 |
| Anxiety | Soiling (Age 9) | 1.08 (0.96-1.22) | 0.18 |
| Anxiety | Soiling (Age 14) | 1.00 (0.87-1.14) | 0.95 |
| Anxiety | Constipation (Age 9) | 0.96 (0.88-1.05) | 0.39 |
| Anxiety | Constipation (Age 14) | 0.99 (0.89-1.10) | 0.81 |
| Depression | Daytime Urinary Incontinence (Age 9) | 1.20 (1.06-1.36) | 0.00 |
| Depression | Daytime Urinary Incontinence (Age 14) | 0.98 (0.83-1.16) | 0.81 |
| Depression | Enuresis (Age 9) | 1.00 (0.91-1.10) | 0.98 |
| Depression | Enuresis (Age 14) | 1.09 (0.90-1.31) | 0.37 |
| Depression | Soiling (Age 9) | 1.08 (0.95-1.22) | 0.23 |
| Depression | Soiling (Age 14) | 0.95 (0.83-1.09) | 0.49 |
| Depression | Constipation (Age 9) | 1.06 (0.97-1.16) | 0.23 |
| Depression | Constipation (Age 14) | 1.06 (0.95-1.18) | 0.30 |

Supplementary Figure 12. Coefficients and 95% confidence intervals for the analysis of PRS Scores at the p < 0.0000001 SNP-inclusion significance level (S12)

| PRS | Incontinence Type | Odds Ratios (95% CI) | *p* |
|---|---|---|---|
| ADHD | Daytime Urinary Incontinence (Age 9) | 1.00 (0.88-1.12) | 0.94 |
| ADHD | Daytime Urinary Incontinence (Age 14) | 0.98 (0.82-1.15) | 0.77 |
| ADHD | Enuresis (Age 9) | 0.96 (0.88-1.06) | 0.43 |
| ADHD | Enuresis (Age 14) | 0.97 (0.81-1.16) | 0.70 |
| ADHD | Soiling (Age 9) | 1.10 (0.97-1.24) | 0.13 |
| ADHD | Soiling (Age 14) | 1.01 (0.88-1.16) | 0.89 |
| ADHD | Constipation (Age 9) | 0.98 (0.90-1.07) | 0.68 |
| ADHD | Constipation (Age 14) | 1.04 (0.93-1.15) | 0.51 |
| Autism | Daytime Urinary Incontinence (Age 9) | 1.04 (0.92-1.17) | 0.52 |
| Autism | Daytime Urinary Incontinence (Age 14) | 1.07 (0.90-1.26) | 0.44 |
| Autism | Enuresis (Age 9) | 1.10 (1.01-1.20) | 0.04 |
| Autism | Enuresis (Age 14) | 1.08 (0.91-1.29) | 0.37 |
| Autism | Soiling (Age 9) | 0.96 (0.85-1.09) | 0.55 |
| Autism | Soiling (Age 14) | 0.96 (0.84-1.11) | 0.61 |
| Autism | Constipation (Age 9) | 0.94 (0.86-1.03) | 0.20 |
| Autism | Constipation (Age 14) | 0.99 (0.89-1.10) | 0.85 |
| Intelligence | Daytime Urinary Incontinence (Age 9) | 1.00 (0.88-1.13) | 0.95 |
| Intelligence | Daytime Urinary Incontinence (Age 14) | 1.21 (1.03-1.43) | 0.02 |
| Intelligence | Enuresis (Age 9) | 0.94 (0.85-1.02) | 0.15 |
| Intelligence | Enuresis (Age 14) | 0.93 (0.77-1.11) | 0.43 |
| Intelligence | Soiling (Age 9) | 1.01 (0.89-1.14) | 0.89 |
| Intelligence | Soiling (Age 14) | 1.15 (1.00-1.33) | 0.04 |
| Intelligence | Constipation (Age 9) | 1.03 (0.94-1.12) | 0.60 |
| Intelligence | Constipation (Age 14) | 1.03 (0.92-1.14) | 0.64 |
| Anxiety | Daytime Urinary Incontinence (Age 9) | 1.04 (0.92-1.17) | 0.53 |
| Anxiety | Daytime Urinary Incontinence (Age 14) | 0.93 (0.79-1.10) | 0.42 |
| Anxiety | Enuresis (Age 9) | 0.98 (0.90-1.08) | 0.73 |
| Anxiety | Enuresis (Age 14) | 1.04 (0.87-1.24) | 0.69 |
| Anxiety | Soiling (Age 9) | 1.10 (0.98-1.24) | 0.11 |
| Anxiety | Soiling (Age 14) | 0.98 (0.85-1.12) | 0.75 |
| Anxiety | Constipation (Age 9) | 1.01 (0.92-1.10) | 0.87 |
| Anxiety | Constipation (Age 14) | 0.92 (0.82-1.02) | 0.12 |
| Depression | Daytime Urinary Incontinence (Age 9) | 1.20 (1.06-1.36) | 0.00 |
| Depression | Daytime Urinary Incontinence (Age 14) | 0.96 (0.81-1.13) | 0.63 |
| Depression | Enuresis (Age 9) | 0.98 (0.90-1.08) | 0.69 |
| Depression | Enuresis (Age 14) | 1.01 (0.84-1.21) | 0.93 |
| Depression | Soiling (Age 9) | 1.10 (0.98-1.24) | 0.12 |
| Depression | Soiling (Age 14) | 0.96 (0.83-1.10) | 0.53 |
| Depression | Constipation (Age 9) | 1.06 (0.97-1.17) | 0.18 |
| Depression | Constipation (Age 14) | 1.07 (0.96-1.19) | 0.22 |

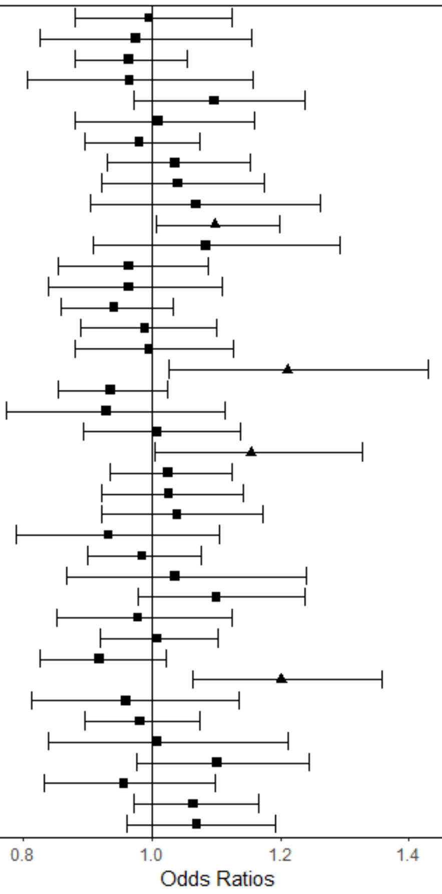

Supplementary Figure 13. Coefficients and 95% confidence intervals for the analysis of PRS Scores at the p < 0.00000005 SNP-inclusion significance level (S13)

| PRS | Incontinence Type | Odds Ratios (95% CI) | *p* |
|---|---|---|---|
| ADHD | Daytime Urinary Incontinence (Age 9) | 1.02 (0.90-1.15) | 0.78 |
| ADHD | Daytime Urinary Incontinence (Age 14) | 1.01 (0.85-1.19) | 0.91 |
| ADHD | Enuresis (Age 9) | 0.96 (0.88-1.06) | 0.44 |
| ADHD | Enuresis (Age 14) | 0.98 (0.82-1.17) | 0.82 |
| ADHD | Soiling (Age 9) | 1.09 (0.96-1.23) | 0.17 |
| ADHD | Soiling (Age 14) | 1.03 (0.90-1.18) | 0.69 |
| ADHD | Constipation (Age 9) | 1.00 (0.91-1.09) | 0.97 |
| ADHD | Constipation (Age 14) | 1.04 (0.94-1.16) | 0.44 |
| Autism | Daytime Urinary Incontinence (Age 9) | 1.04 (0.92-1.17) | 0.52 |
| Autism | Daytime Urinary Incontinence (Age 14) | 1.07 (0.90-1.26) | 0.44 |
| Autism | Enuresis (Age 9) | 1.10 (1.01-1.20) | 0.04 |
| Autism | Enuresis (Age 14) | 1.08 (0.91-1.29) | 0.37 |
| Autism | Soiling (Age 9) | 0.96 (0.85-1.09) | 0.55 |
| Autism | Soiling (Age 14) | 0.96 (0.84-1.11) | 0.61 |
| Autism | Constipation (Age 9) | 0.94 (0.86-1.03) | 0.20 |
| Autism | Constipation (Age 14) | 0.99 (0.89-1.10) | 0.85 |
| Intelligence | Daytime Urinary Incontinence (Age 9) | 0.99 (0.87-1.12) | 0.84 |
| Intelligence | Daytime Urinary Incontinence (Age 14) | 1.21 (1.03-1.43) | 0.02 |
| Intelligence | Enuresis (Age 9) | 0.94 (0.86-1.03) | 0.19 |
| Intelligence | Enuresis (Age 14) | 0.96 (0.80-1.15) | 0.63 |
| Intelligence | Soiling (Age 9) | 1.02 (0.91-1.15) | 0.73 |
| Intelligence | Soiling (Age 14) | 1.15 (1.00-1.32) | 0.05 |
| Intelligence | Constipation (Age 9) | 1.05 (0.96-1.15) | 0.29 |
| Intelligence | Constipation (Age 14) | 1.04 (0.93-1.16) | 0.49 |
| Anxiety | Daytime Urinary Incontinence (Age 9) | 1.04 (0.92-1.17) | 0.53 |
| Anxiety | Daytime Urinary Incontinence (Age 14) | 0.93 (0.79-1.10) | 0.42 |
| Anxiety | Enuresis (Age 9) | 0.98 (0.90-1.08) | 0.73 |
| Anxiety | Enuresis (Age 14) | 1.04 (0.87-1.24) | 0.69 |
| Anxiety | Soiling (Age 9) | 1.10 (0.98-1.24) | 0.11 |
| Anxiety | Soiling (Age 14) | 0.98 (0.85-1.12) | 0.75 |
| Anxiety | Constipation (Age 9) | 1.01 (0.92-1.10) | 0.87 |
| Anxiety | Constipation (Age 14) | 0.92 (0.82-1.02) | 0.12 |
| Depression | Daytime Urinary Incontinence (Age 9) | 1.19 (1.05-1.35) | 0.01 |
| Depression | Daytime Urinary Incontinence (Age 14) | 0.95 (0.80-1.12) | 0.54 |
| Depression | Enuresis (Age 9) | 0.97 (0.89-1.06) | 0.54 |
| Depression | Enuresis (Age 14) | 0.99 (0.82-1.19) | 0.89 |
| Depression | Soiling (Age 9) | 1.09 (0.96-1.23) | 0.18 |
| Depression | Soiling (Age 14) | 0.98 (0.85-1.12) | 0.73 |
| Depression | Constipation (Age 9) | 1.06 (0.96-1.16) | 0.24 |
| Depression | Constipation (Age 14) | 1.03 (0.93-1.15) | 0.58 |

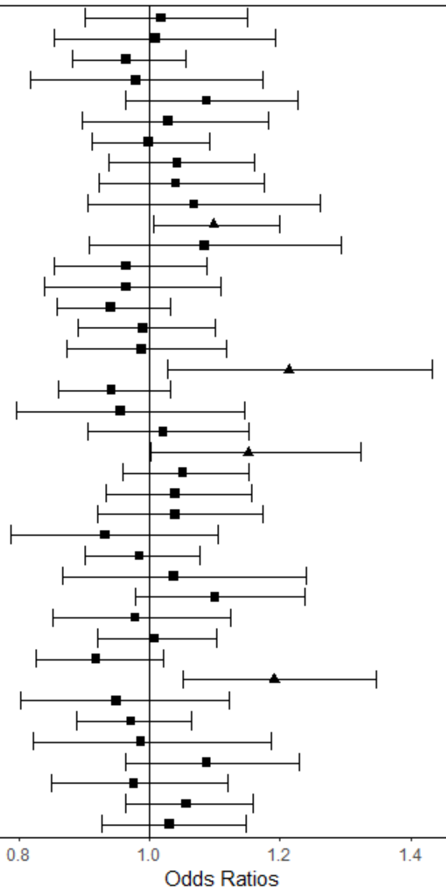
